# Automated echocardiography image quality assessment and its association with AI performance

**DOI:** 10.64898/2026.11.19.26368489

**Authors:** Pere Lopez-Gutierrez, Juan Garrido-Oliver, Mireia Bragulat-Arevalo, Juan Pablo Betancur-Rengifo, Nicholas Cauwenberghs, Alberto Morales-Galan, Marawan Elbatel, Machteld J. Boonstra, Francesca Ferrando, Horacio Majul, Lisandro Rivas Catoni, Santiago Martin Grieve, Stephan van der Zwaard, Yi Qin, Folkert W. Asselbergs, Tatiana Kuznetsova, Gisela Teixido-Tura, Alex P.W. Lee, Xiaomeng Li, Karim Lekadir, Laura Galian-Gay, Jose Rodriguez-Palomares, Ignacio Ferreira-Gonzalez, Lydia Dux-Santoy, Andrea Guala

**Affiliations:** Vall d’Hebron Institut de Recerca, Barcelona, Spain; Universitat Autònoma de Barcelona, Bellaterra, Spain; KU Leuven, Leuven, Belgium; The Hong Kong University of Science and Technology, Hong Kong SAR, China; Amsterdam University Medical Center, University of Amsterdam, Amsterdam, the Netherlands; CIBER-CV, Instituto de Salud Carlos III, Madrid, Spain; University College London, London, United Kingdom; The Chinese University of Hong Kong (Department of Medicine & Therapeutics), Hong Kong, China; Institució Catalana de Recerca i Estudis Avançats (ICREA); Artificial Intelligence in Medicine Lab (BCN-AIM), Universitat de Barcelona, Spain; Centro de Investigación en Red de Epidemiología y Salud Pública (CIBERESP), Madrid, Spain

**Keywords:** echocardiography, image quality, artificial intelligence, deep learning

## Abstract

**Background:** Echocardiographic image quality influences clinical interpretation, yet its definition remains subjective and its impact on artificial intelligence (AI) systems has not been systematically evaluated. We developed EchoQuality, a deep-learning model for automated echocardiography quality assessment, and quantified the effect of image quality on AI performance across a broad range of tasks.

**Methods:** A multidisciplinary co-creation process established a definition of echocardiographic image quality, which was used to annotate 6,474 videos for EchoQuality development and internal testing, and 1,000 videos for external testing. The impact of image quality on the performance of state-of-the-art AI models was subsequently assessed for the prediction of 12 measurements, and the detection of 5 moderate dysfunctions and 6 cardiac diseases across five international cohorts totalling 762,588 videos from 25,205 echocardiography studies. In addition, a quality-aware aggregation strategy was developed to mitigate the impact of low-quality images.

**Results:** EchoQuality achieved a performance comparable to the inter-observer agreement, while the agreement with an expert in a prospective, blinded best-clip selection experiment was 90.4%. In downstream AI uses, high-quality studies resulted in the highest performance in 97 of 122 (80%) of evaluations. The median performance gaps between high- and low-quality videos were 10% [5-20%] in the prediction of quantitative measurements (R^2^), 14% [7-20%] in the identification of moderate dysfunctions (sensitivity), 12% [3-20%] in the detection of diseases (sensitivity), 12% [9-15%] in view identification (accuracy), and 3% [2-5%] in left-ventricular segmentation (Dice score). Performance gaps were consistent across state-of-the-art models, datasets, and tasks. Finally, a quality-aware aggregation strategy improved performance in 88% (63 of 72) of the evaluations, with a median relative improvement of 5% [1-15%].

**Conclusions:** EchoQuality provides an automated assessment of echocardiographic image quality, a major determinant of AI performance. Incorporating quality-awareness may improve the robustness and clinical reliability of echocardiographic AI systems.

## Introduction

Echocardiography is a pillar of clinical cardiology, being indicated throughout the spectrum of practice. Yet, the technology and manual acquisition of echocardiographic images entails high variability in image appearance and quality. Echocardiography image quality is assessed visually to estimate the degree of uncertainty of eventual clinical findings^1,2^, and it has been shown to depend on patient body habitus, clinical condition, patient position, sonographer experience, and scanner^3–5^, among others. Importantly, limited image quality negatively impacts the accuracy of the standard quantification of left ventricular (LV) ejection fraction^6–8^, size^5^, mass^9^, strain^5,10,11^, and other features^12^.

Artificial intelligence (AI) applications to echocardiography have been growing. Examples include the support of acquisition^13–15^, view identification^16,17^, and the assessment of multiple descriptors and diagnoses^17–21^. Most of research in automatic image quality assessment in echocardiography focused on quality control for AI-guided image acquisition^13–15,22,23^ and mostly defined “quality” as the extent to which a given video corresponds to one of the standard imaging planes. Beyond applications related to AI-guided acquisition, several works presented AI models to predict echocardiographic image quality^24–29^. Nonetheless, most of image quality definitions included a mix of view-related characteristics, such as foreshortening and the presence of the expected cardiac structures, and image appearance characteristics. Only two previous works presented models aiming at predicting image quality beyond structural and view-related characteristics^28^. Among them, one article presented an external validation^28^, none a prospective validation.

The impact of predicted image quality on the performance of AI models in downstream tasks was included in two works^24,28^, both limited to left-ventricular ejection fraction assessment, and no paper reported the impact on the performance of state-of-the-art AI models for echocardiography, nor in the prediction of cardiac diseases. Thus, whether echocardiography image quality affects AI models performance remains largely unexplored.

We hypothesized that an AI model, hereinafter referred to as EchoQuality, could replicate human-based quality assessment, permitting reproducible and scalable image quality grading. We further hypothesized that image quality systematically impacts AI model performance and that incorporating awareness of image quality into AI inference can improve accuracy.

## Methods

### Data

Six cohorts were analyzed (Figure 1). Firstly, Hospital Universitari Vall d’Hebron (HUVH), the EchoQuality development center, contributed a clinical dataset comprising 15,063 transthoracic echocardiographic (TTE) studies from 9,841 patients acquired during routine clinical care between 2019 and 2023. The local Ethics Committee waived informed consent. This dataset was randomly split at patient level into three non-overlapping datasets: (i) HUVH-quality, comprising 8,840 videos from 463 studies from 458 patients, of which 6,474 were retained after excluding videos classified as other views, further randomly split into 5,056 training and 1,418 testing videos, the latter constituting also the HUVH-view dataset used for view identification evaluation; (ii) HUVH-testing, including 442,090 videos from 14,500 studies from 9,283 patients to test the impact of image quality in AI models performance; and (iii) HUVH-BestClip, a set of 100 studies of 100 patients to be prospectively analyzed (see below). Additionally, a dedicated research dataset (HUVH-POCUS) comprising 2,458 conventional TTE videos and 1,521 same-day point-of-care ultrasound (POCUS) videos from 100 consecutive patients, each signing informed consent, was available.

**Figure 1.**
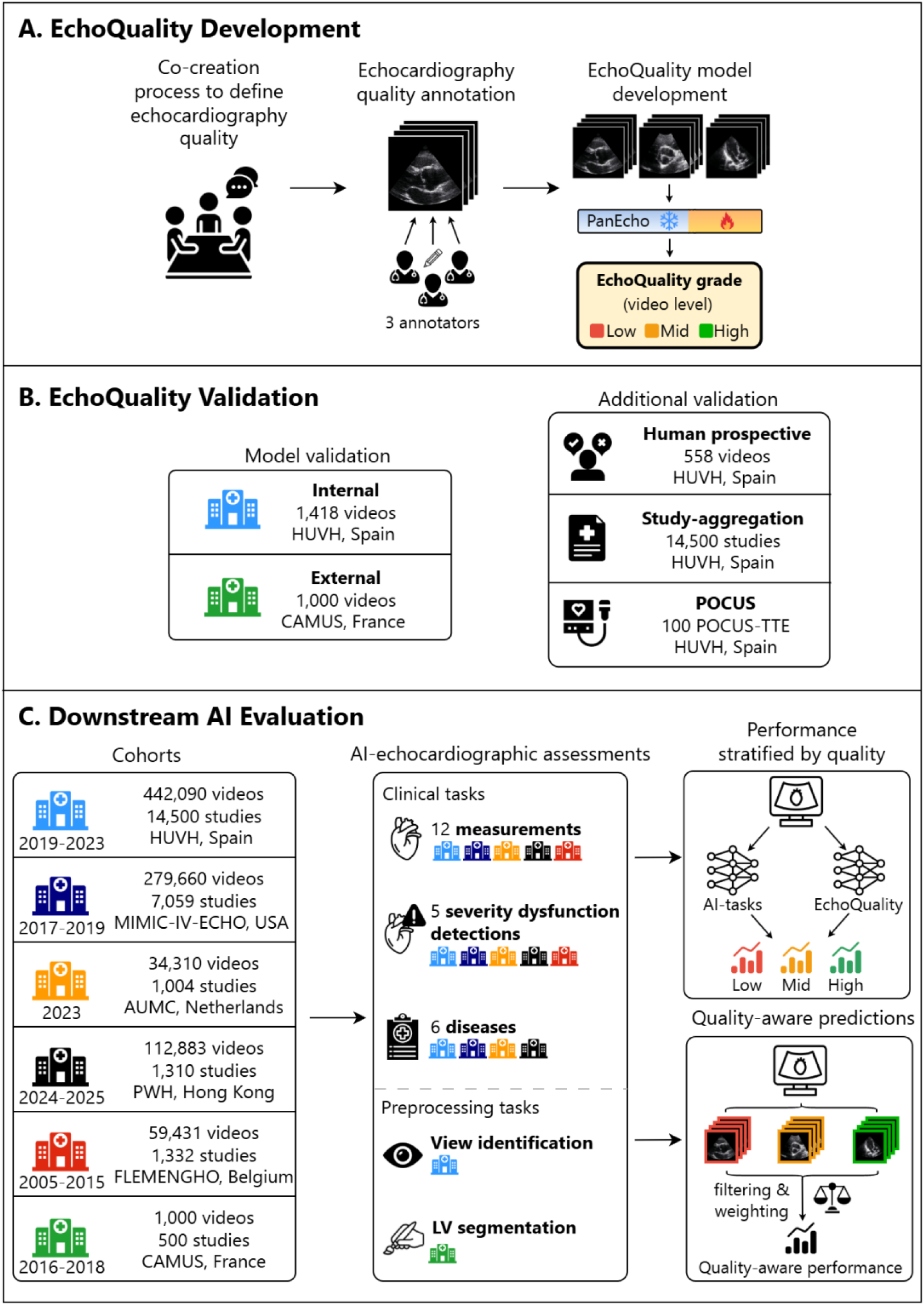
Overview of the development, validation, and downstream evaluation of EchoQuality. (A) Development through expert-defined quality criteria, annotations, and model training. (B) Internal and external validations. (C) Evaluation of the impact of image quality on downstream AI tasks and quality-aware aggregation.

Five external datasets were used to externally validate EchoQuality predictions or to assess image quality impact in AI performance. One was the (i) MIMIC-IV-ECHO^30–34^, including 279,660 videos from 7,059 studies from 4,552 patients in the United States. Following^35^, measurements were extracted from videos using optical character recognition (Figure S1). The (ii) Prince of Wales Hospital (PWH, Hong Kong) provided 112,883 videos from 1,310 studies acquired in 1,238 patients during clinical practice, while the (iii) Amsterdam University Medical Center (AUMC, The Netherlands) contributed 34,310 videos from 1,004 studies acquired in 890 patients during clinical practice. The (iv) Flemish Study on Environment, Genes and Health Outcomes (FLEMENGHO, Belgium), a population-based cohort study, provided 59,431 videos from 1,332 studies and 1,332 individuals. Finally, the (v) CAMUS^36^ dataset (France), contributed 1,000 apical 2- and 4-chamber videos from 500 patients.

### EchoQuality development

#### Image quality definition

During a co-creation process involving cardiologists, sonographers, and researchers, the key characteristics determining the perception of image quality were discussed (Figure 1A). The discussion produced an initial discrimination between characteristics related to the imaging view and those related to image appearance. Regarding view-related characteristics, the participants agreed on the importance of (i) acquiring standard views, (ii) the presence of all structures expected in that view, and, in apical views, (iii) the correct alignment with left-ventricular (LV) long axis. Regarding image appearance, participants agreed that the key characteristic is (iv) the difference in image brightness between cardiac structures and blood pool, or, in other words, the contrast between blood and cardiac structures, which determines structural boundaries definition. As well-performing, openly-available AI algorithms for view identification can identify suboptimal views^16,17^, subsequent model development focused on the image-appearance characteristic. This metric, hereinafter referred to as image quality, was selected as the primary quality metric to be predicted by EchoQuality and was categorized using a three-level scale as “low”, “mid”, and “high”.

#### Annotations

Following co-creation, three experienced cardiologists annotated the HUVH-quality dataset. The annotation included (i) view (apical 2-, 3-, 4- and 5-chamber, parasternal long- and short-axis, and “other views”), (ii) presence of all expected cardiac structures, (iii) apical foreshortening, and (iv) image quality. A subset of 350 videos was independently annotated by two experts to assess inter-observer reproducibility. Furthermore, a cardiologist annotated all 1000 CAMUS videos, hereinafter called “in-house” annotations to distinguish them from the “original” quality annotations of CAMUS. In addition, in the HUVH-BestClip dataset, an independent clinician not involved in the co-creation sessions prospectively identified the best video per view, to be compared with EchoQuality grading in the prospective validation.

#### Model development and validation

EchoQuality was developed as a deep learning model to assess the quality of 2D B-mode echocardiography videos. The model was trained by fine-tuning PanEcho^19^ with the HUVH-quality dataset, using view classification, the three above-mentioned view-related characteristics, and image quality as video-level multitask outputs. Additional implementation details are provided in Tables S1 and S2. Model weights are available at https://github.com/CardiovascularImagingVallHebron/EchoQuality.

For clinical tasks with study-level outcome a study-level quality metric was required. Mirroring how variable quality is handled in clinical practice, video-level EchoQuality predictions were aggregated using a two-stage hierarchical rule: first, within each view, the highest-quality video defined the view-level quality grade; second, view-level grades were aggregated to study level using majority voting, with highest quality winning in case of draw.

Finally, to evaluate the effect of acquisition modalities on image quality, EchoQuality grade was computed separately for conventional and POCUS videos in the HUVH-POCUS dataset.

### Downstream evaluation

To characterize the impact of image quality on AI performance, EchoQuality was deployed in a large-scale, multicenter inference spanning heterogeneous cohorts, AI models, and echocardiographic assessments. Three classes of clinical tasks and two standard AI pre-processing tasks were considered.

#### Clinical tasks

Two state-of-the-art AI models, PanEcho and EchoPrime^17^, were evaluated for 23 predictions, grouped in three sets of tasks.

The first set of tasks concerned the quantification of twelve measurements: LV ejection fraction (LVEF), LV end-diastolic and end-systolic volumes (LVEDV, LVESV), LV diastolic and systolic diameters, LV posterior wall and interventricular septal thickness, left atrial (LA) end-systolic volume and diameter, mitral inflow velocity ratio (E/e’), tricuspid systolic excursion (TAPSE), and aortic valve peak velocity (AV Vmax). These analyses were performed in all but CAMUS datasets, and center- and quality-specific results were reported if at least 50 observations were available.

The second set of tasks concerned the detection of five at least moderate dysfunctions: LV systolic dysfunction (defined as LVEF<40%), aortic valve stenosis (Vmax≥3 m/s), LV dilation (indexed LVEDV>90 ml/m^2^ in males, >71 in females), LV hypertrophy (LV wall thickness≥14 mm in males, ≥13 in females), and LA dilation (indexed LA volume≥42 mL/m^2^). These analyses were performed in all but CAMUS datasets, and center- and quality-specific results were reported if at least 20 positive cases were available.

The third set of tasks concerned the detection of six diseases: aortic valve regurgitation and stenosis, mitral valve regurgitation and stenosis, pericardial effusion, and tricuspid valve regurgitation. These analyses were performed in four datasets, as FLEMENGHO and CAMUS have no labels, and center- and quality-specific results were reported if at least 20 positive cases were available.

#### Preprocessing tasks

For basic AI-preprocessing tasks, view identification was evaluated using the Zhang model^16^ and EchoPrime^17^ in the HUVH-view dataset. LV segmentation was evaluated using two models^37–39^ trained on EchoNetDynamic^40^ and CAMUS, and tested on CAMUS.

### Quality-aware aggregation

To test whether image quality could be exploited to improve study-level predictions, we implemented a quality-aware strategy designed to emulate how video-level information is aggregated in echocardiographic practice, where measurements derived from suboptimal images are disregarded in favor of those obtained from high-quality images. Accordingly, video-level predictions were aggregated using a continuous EchoQuality-weight, such that higher-quality videos contribute more to study-level predictions (Supplementary Material). This analysis was restricted to measurements and moderate dysfunctions detection using PanEcho, as it provides video-level outputs suitable for aggregation.

### Statistical analysis

Balanced accuracy was used to assess inter-observer agreement and EchoQuality validation performance, the latter additionally complemented by area under the ROC curve (AUROC). In downstream assessments, performance was evaluated using R² and mean absolute error (MAE) for measurements, sensitivity and specificity for moderate dysfunctions and disease detection, accuracy for view identification, and Dice score for LV segmentation. For disease detection, cohort-specific optimal thresholds were derived via Youden index. For all analyses, 95% confidence intervals were computed using 1,000 bootstraps. For PanEcho analysis video-level predictions were aggregated to study level using only the views where the structure to be measured is visible while for the Zhang view identification model grouping of view predictions was performed (Table S3 and S4). Group comparisons were performed using the Kruskal-Wallis or Mann–Whitney tests for continuous variables and the chi-square test for categorical variables. All analyses were performed using Python 3.11.9, and a p-value≤0.050 was considered significant.

## Results

Results are presented in three sections: first, EchoQuality validation, second, impact of image quality on downstream AI performance, and finally the impact of quality-aware predictions.

### EchoQuality validation

In the internal testing set of the HUVH-quality dataset, which includes 630 (40%) low-, 663 (47%) mid-, and 125 (9%) high-quality videos, EchoQuality achieved a balanced accuracy of 63.2% [CI95%: 59.8-66.0%], comparable to the inter-observer variability 47.9% [38.3-58.5%]. The discriminative performance was good (AUROC 0.784 [0.778–0.790]), with best separation for high-quality videos (0.863 [0.852–0.875]) and lower for mid-quality videos (0.658 [0.647–0.667]), reflecting the intrinsic ambiguity of intermediate quality. In the external validation cohort, EchoQuality achieved a balanced accuracy of 50.9% [44.4-57.1%] and overall AUROC of 0.681 [0.658–0.703] against the in-house annotations, and a balanced accuracy of 52.3% [49.4-55.1%] and an AUROC of 0.651 [0.628-0.676] against original CAMUS quality grades. Of note, the agreement between in-house and original CAMUS annotations was very limited (21.0% [18.5-23.5%]), with in-house annotations systematically assigning lower quality (Figure S2).

In the prospective validation study on the HUVH-BestClip cohort, expert clip selection was strongly aligned with EchoQuality grade ranking: the expert selected a video from the highest EchoQuality grade in 90.4% (207/229) of cases.

Regarding the aggregation of video-level predictions into study-level quality grades, aggregated EchoQuality grades showed strong concordance with quality annotations from echocardiographic reports (Figure 2A). For example, 67% of studies described as “bad quality” during clinical care were classified as low-quality by EchoQuality, whereas 12% of studies described as good or excellent during care were classified as low quality by EchoQuality.

**Figure 2.**
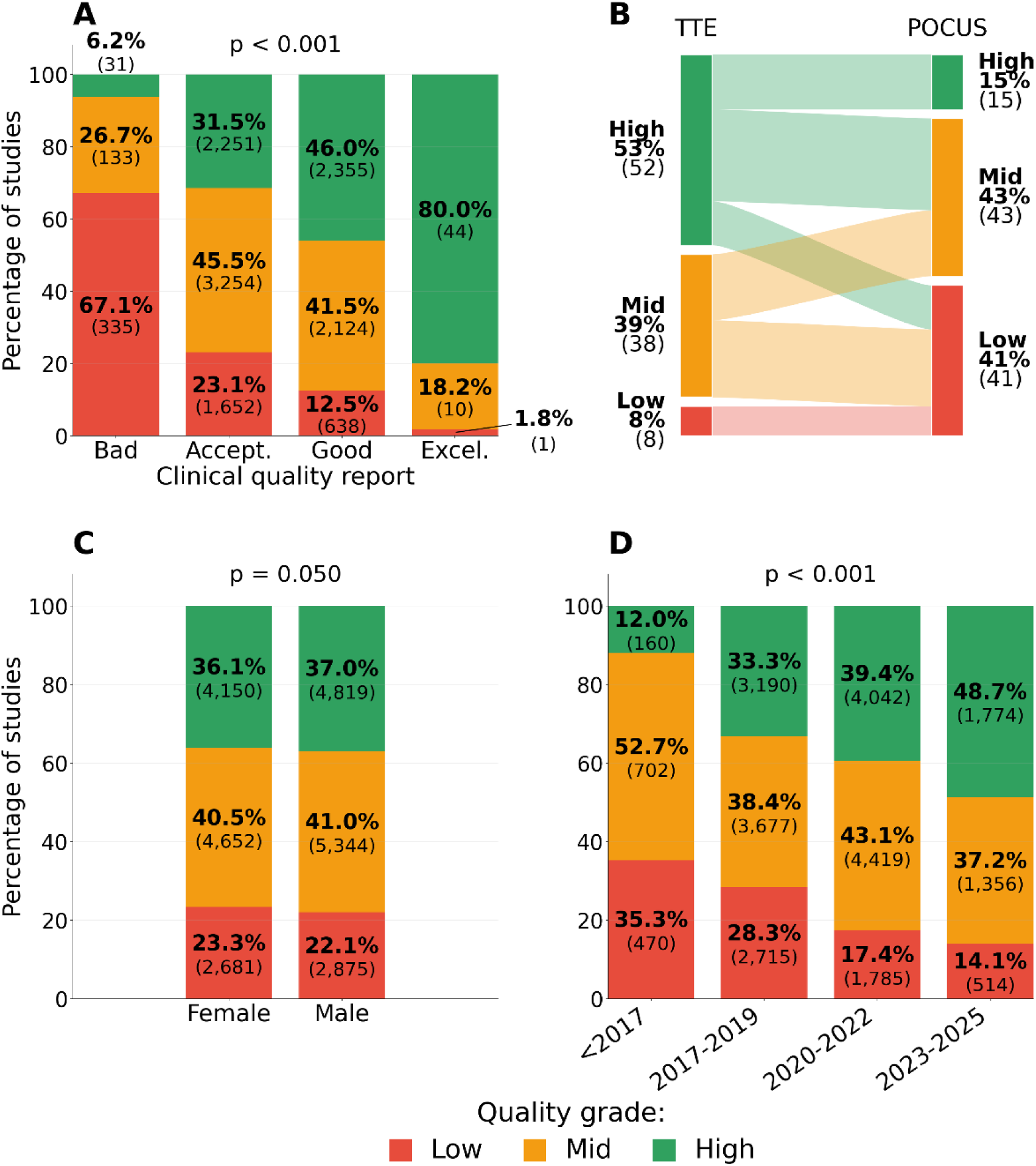
(A) Distribution of study-level EchoQuality by clinical quality assessment. (B) Comparison of EchoQuality study-level grades between same-day conventional echocardiography and POCUS examinations. Differences in EchoQuality according to (C) patient sex, and (D) year of acquisition.

In the HUVH-POCUS dataset, EchoQuality grades were consistently higher in conventional studies (52 high, 39 mid, 8 low) compared with same-day POCUS (15 high, 43 mid, 41 low), with no patient showing lower quality in conventional echocardiography than in POCUS (Figure 2B).

### Impact of image quality

A total of 762,588 videos from 25,205 echocardiography studies across five cohorts and three continents were included (Table 1).

**Table 1.**
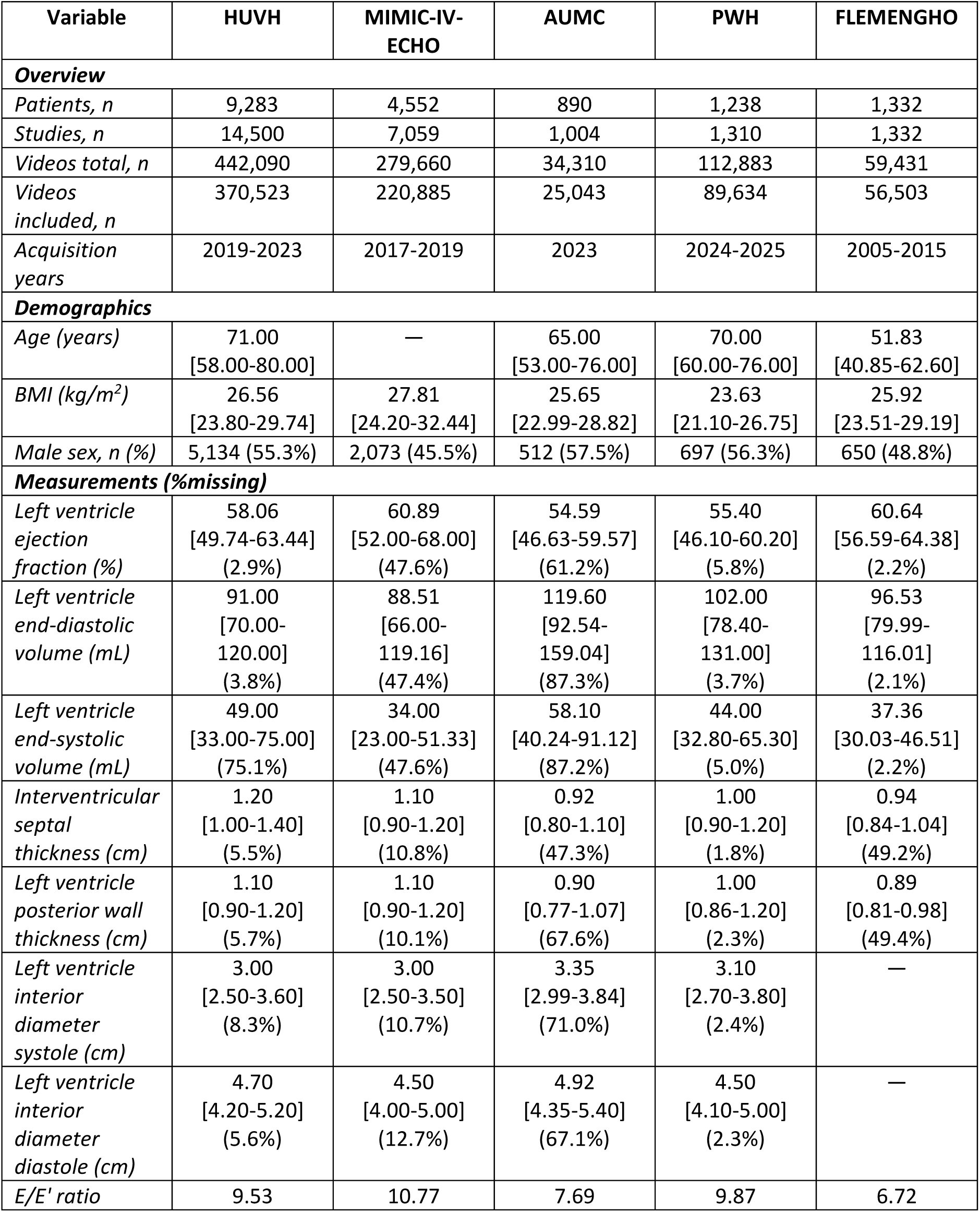
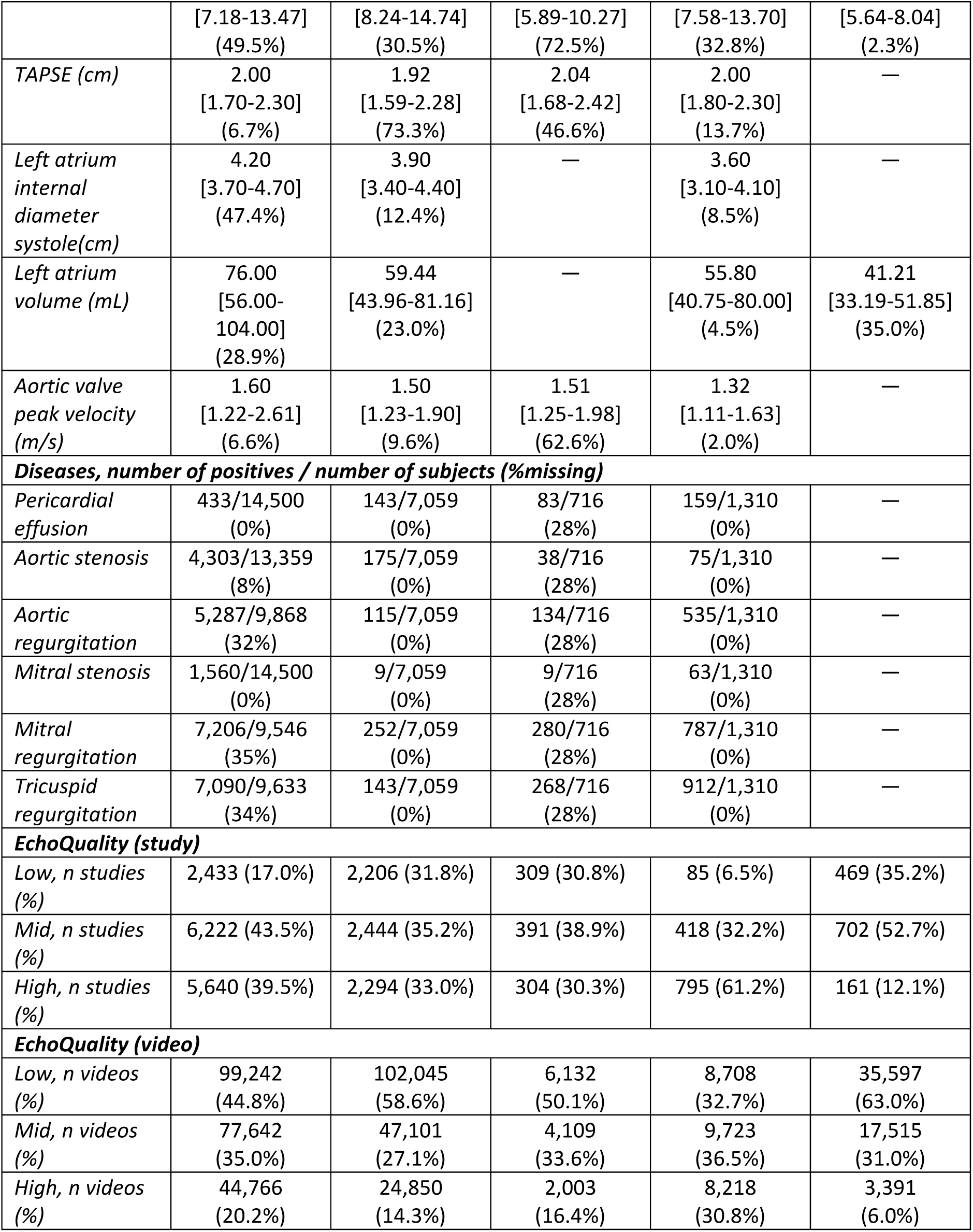
Cohort overview.

#### Clinical, demographic, and temporal patterns

Age was inversely associated with EchoQuality grade in HUVH-testing, AUMC and FLEMENGHO (p < 0.05) but not in PWH (p = 0.242), where the age range was minor, while minimal differences were noted with respect to patient sex (p = 0.050, Figure 2C). BMI was inversely associated with EchoQuality grade in every cohort (all p < 0.050). Over time, the overall proportion of low-quality studies decreased from 35.3% before 2017 to 14.1% by 2023-2025, while the proportion of high-quality studies increased from 12.0% to 48.7% (Figure 2D). This temporal analysis was replicated in the HUVH cohort, where the same monotonic improvement in quality distribution over time was observed (Figure S3).

#### AI-based assessments

Across all measurements, severity stratifications, disease detections, preprocessing tasks, and cohorts and models, high-quality studies achieved the best performance in 97 of 122 evaluations (80%), compared with 16 (13%) for mid-and 9 (7%) for low-quality studies. In the HUVH-testing cohort, where study-level quality was visually graded during care, the differences in performance of AI models with respect to image quality were equivalent to those obtained with EchoQuality (Figure S4, Table S5).

#### Measurements

High-quality studies achieved the best performance in 42 of 56 measurements (75%), with R² difference between high-and low-quality studies of 0.10 [0.05-0.20] (Figure 3). R² values across all cohorts and measurements were 0.47 [0.27-0.59], 0.36 [0.27-0.50], and 0.33 [0.15-0.46] for high-, mid-, and low-quality studies, respectively. Cohort-specific median R² in high-, mid-, and low-quality videos were: HUVH 0.54, 0.47, 0.40; MIMIC-IV-ECHO 0.50, 0.38, 0.33; PWH 0.45, 0.36, 0.33; AUMC 0.51, 0.33, 0.29; and FLEMENGHO 0.18, 0.20, 0.06. These performance differences were confirmed for MAE (Figure S4), and were consistent across the vast majority of measurements, and the two models (Table S6). The quality-dependent effect was particularly evident for LVEF, where the mean R² gap between high-and low-quality studies across cohorts was 0.26 for PanEcho and 0.23 for EchoPrime. Importantly, across measurements, cohorts, and models, predictions in low-quality studies showed a more marked tendency to regress toward the cohort-specific median values compared with predictions in mid-and high-quality studies, as illustrated for LVEF in Figure S6.

**Figure 3.**
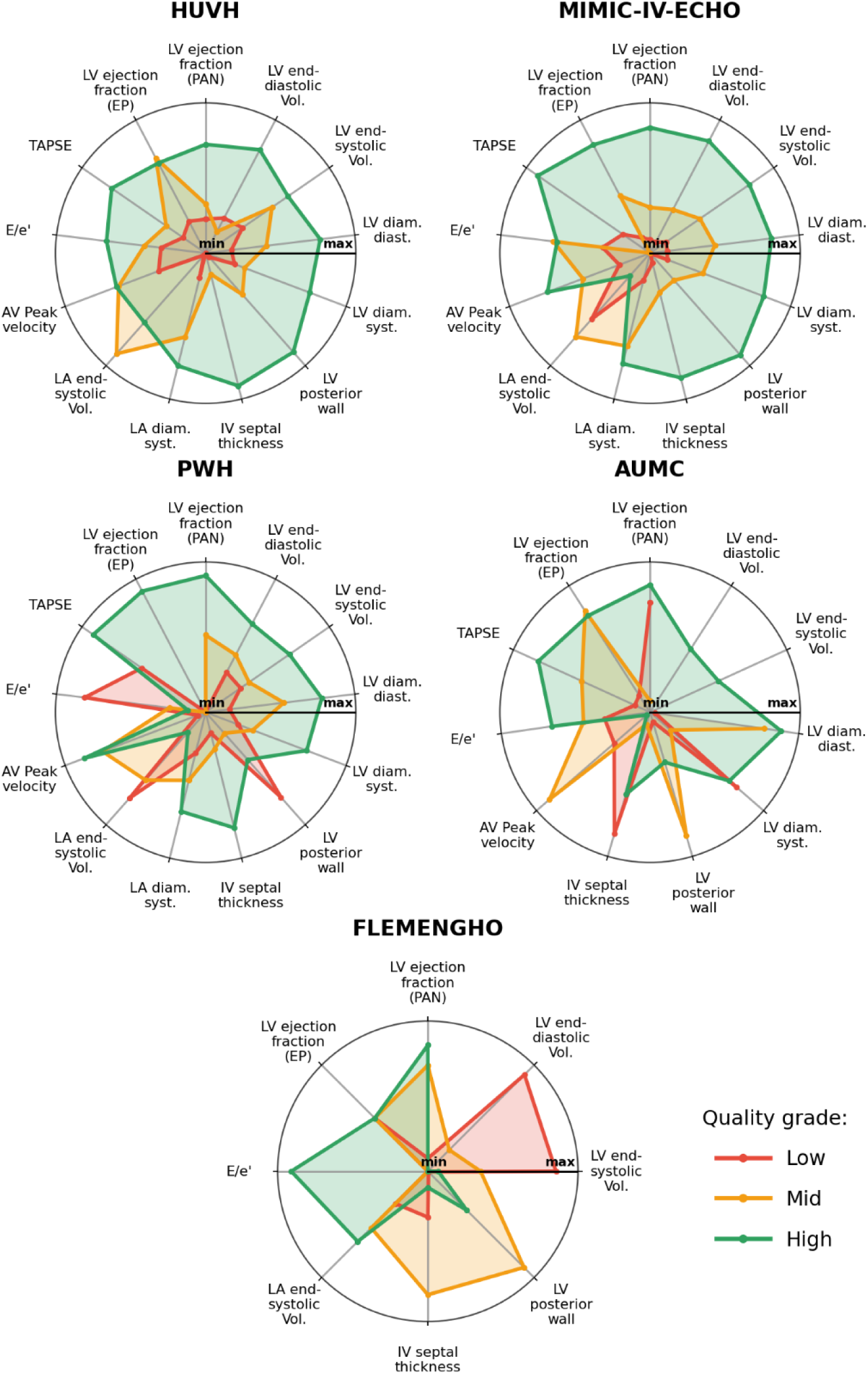
Performance of PanEcho (PAN) and EchoPrime (EP) in the prediction of twelve echocardiographic measurements stratified by EchoQuality grades across five cohorts. Performance is reported as R² values with axis limits set to 0.9 times the minimum R^2^ and 1.1 times the maximum R^2^. Abbreviations: IV = intraventricular, Vol. = volume, Diam. = diameter, Diast. = diastolic, Syst. = systolic.

#### Detection of moderate dysfunctions

The detection of at least moderate dysfunctions showed large differences in AI performances comparing quality grades, with high-quality studies resulting in the best performance in 17 of 20 cases (85%), compared with 3 (15%) and 0 (0%) for mid-and low-quality studies, respectively. The median sensitivity difference between high-and low-quality studies was 0.14 [0.07-0.20]. Sensitivities across all cohorts and dysfunction types were 0.39 [0.19-0.52], 0.31 [0.14-0.35], and 0.26 [0.09-0.31] in high-, mid-, and low-quality studies, respectively. Cohort-specific sensitivities in high-, mid-, and low-quality videos were: HUVH 0.42, 0.34, 0.32; MIMIC-IV-ECHO 0.22, 0.10, 0.11; PWH 0.38, 0.32, N/A; AUMC 0.22, N/A, N/A, respectively, where N/A indicates quality strata with too few positive cases. Notably, this quality-dependent difference in sensitivity was consistently observed across cohorts, conditions, and models (Figure 4, Table S7), and was accompanied by very modest increase in specificity, which remained high across quality strata. The differences were particularly pronounced for the detection of LV systolic dysfunction, LV dilation, and LA dilation, where the mean sensitivity gap between high-and low-quality studies were 27%, 21%, and 17%, respectively. Remarkably, models more frequently failed to identify positive cases in low-quality compared with high-quality studies, favoring the majority class.

**Figure 4.**
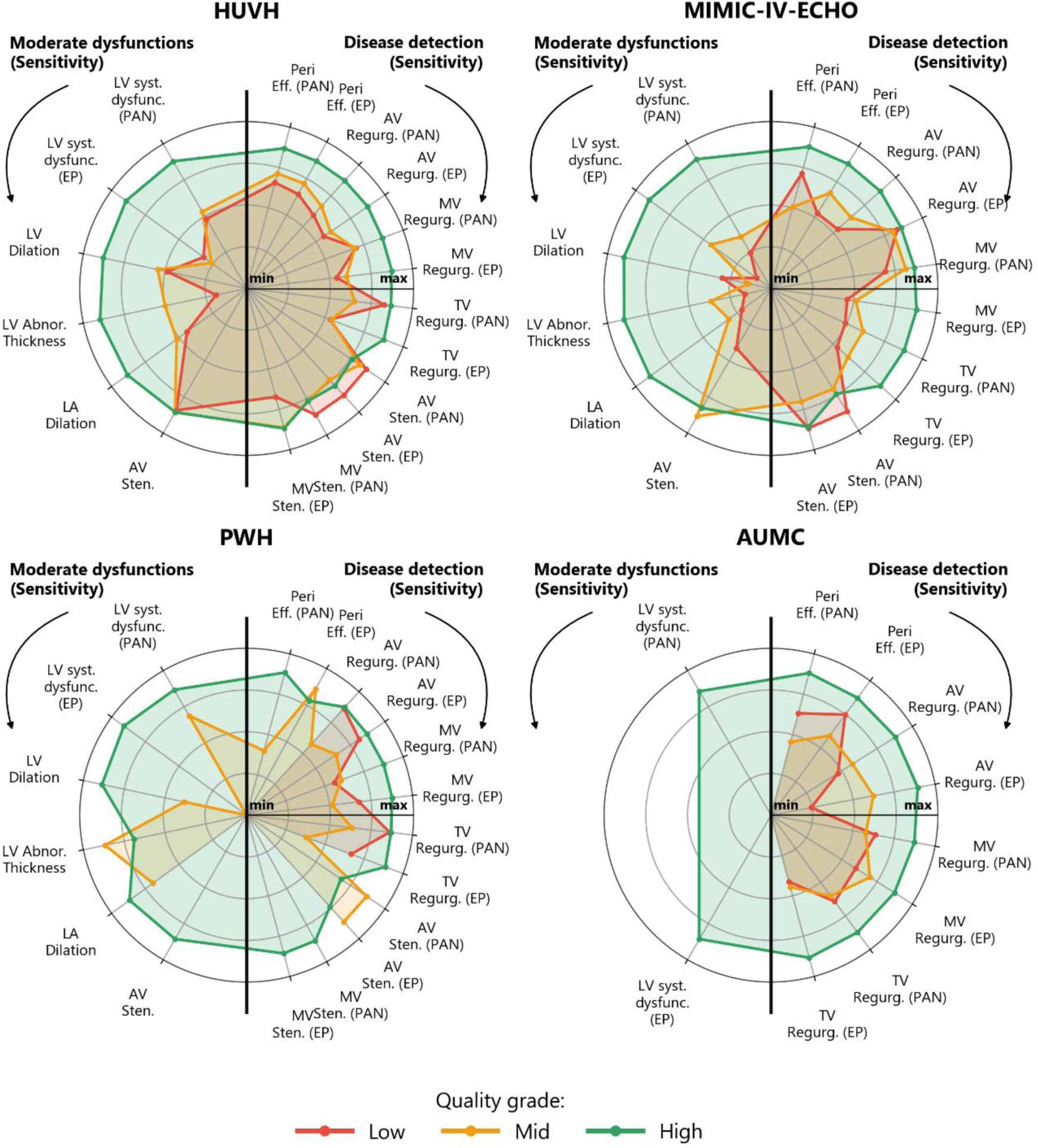
Performance of PanEcho (PAN) and EchoPrime (EP) on moderate dysfunction detection and disease classification tasks stratified by EchoQuality grade. The left half of each radar plot shows sensitivity for moderate dysfunction detection, while the right half shows it for disease classification. For visualization purposes, axis limits were normalized independently for each task and cohort. Abbreviations: AV, MV, TV = aortic, mitral and tricuspid valve, respectively, Sten. = stenosis, Regurg. = regurgitation, Abnor. = abnormal, Syst. = systolic, Dysf = dysfunction, Peri. Eff. = pericardial effusion.

#### Disease detection

Across the four datasets with available diagnostic information, high-quality studies obtained the best performance in 34 of 42 evaluations (81%), compared with 3 (7%) and 5 (12%) of cases in mid-and low-quality studies, respectively. The median difference in sensitivity in high-quality compared with low-quality studies was 0.12 [0.03-0.20], with sensitivity of 0.75 [0.59-0.83], 0.60 [0.50-0.69], and 0.61 [0.50-0.70] in high-, mid-, and low-quality studies, respectively. Cohort-specific AUROC in high-, mid-, and low-quality videos were: HUVH 0.73, 0.61, 0.62; MIMIC-IV-ECHO 0.79, 0.66, 0.63; PWH 0.78, 0.58, 0.64; AUMC 0.63, 0.43, 0.49. This behavior was consistent over diseases, cohorts, and models (Figure 4, Table S8), and was accompanied by moderate negative differences of −0.06 [-0.14—0.03] in specificity.

#### Preprocessing tasks

In a total of 1,418 videos from the HUVH-view dataset, the accuracy of both models, i.e., Zhang and EchoPrime, in view identification was consistently associated with EchoQuality grade. In particular, the accuracy obtained by the Zhang model increased from 82.5% in low-quality to 92.3% in mid-and 95.8% in high-quality videos, while for EchoPrime accuracies were 93.6, 96.9% and 98.4%, respectively, with consistent trends across views (Figure S7). Inter-model agreement in view classification was strongly influenced by image quality: disagreement rates were highest in low-quality videos (21.9%) and much lower in mid-and high-quality videos (10.7% and 6.5%, respectively). Finally, in CAMUS testing set, the Dice score for LV segmentation improved significantly with EchoQuality grade for both models. Absolute increases from low-to high-quality frames were 0.05 for the EchoNetDynamic-trained model (0.75 to 0.80) and 0.02 for the CAMUS-trained model (0.93 to 0.95).

### Quality-aware aggregation

Quality-aware aggregation improved performance in 63 of 72 evaluations (88%), with a relative improvement of 5% [1-15%] (Figure 5, Figure S8, Table S9). The largest gains were observed in the detection of moderate dysfunctions, with an overall sensitivity improvement of 18% [8-23%]. Such improvement was consistent across cohorts: HUVH 18% [7-21%]; MIMIC-IV-ECHO 20% [19-38%]; PWH 14% [6-15%]; AUMC 18% [14-23%]. Similarly, the performances in quantitative measurements showed an overall median R^2^ improvement of 4% [1-6%], which was consistent across cohorts: HUVH 3% [1-5%]; MIMIC-IV-ECHO 5% [3-9%]; PWH 3% [1-6%]; AUMC 3% [0-12%]; FLEMENGHO 1% [-10-40%]. Notably, quality-aware aggregation partially mitigated the tendency of low-quality videos to drive predictions toward cohort-specific central values or more prevalent classes, thereby improving the identification of more extreme values and detection of moderate dysfunctions (Figure S9).

**Figure 5.**
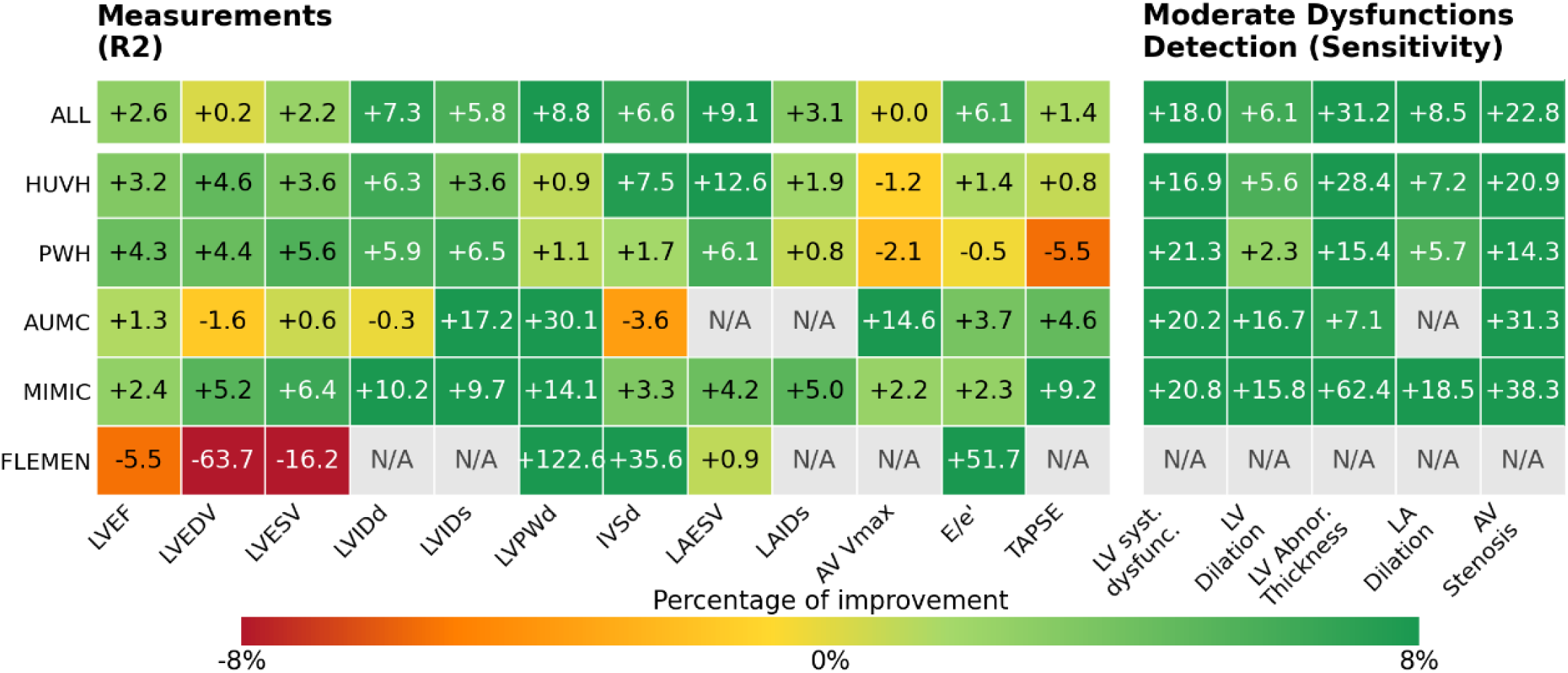
Percentual performance difference comparing standard with quality-aware aggregation. N/A refers to tests whose number of observations was not large enough to compute robust statistics or whose labels were not available.

## Discussion

This study presents EchoQuality, a deep learning model for echocardiographic image quality assessment, and reports the impact of image quality on state-of-the-art echocardiographic AI models in the largest external evaluation ever reported. Through large-scale evaluation across cohorts, models, and prediction tasks, EchoQuality demonstrated consistent stratification of AI performance. Results demonstrated that EchoQuality can be deployed at frame, video, and study levels and enables quality-aware handling of echocardiography videos.

EchoQuality achieved a balanced accuracy of 63% on the internal testing set, and 51 and 52% on the external testing set using in-house and original annotations, respectively, slightly above the 48% inter-observer agreement among cardiologists, underscoring that performance is bound by the inherent subjectivity of quality. This subjectivity was underlined by the limited agreement between the original CAMUS quality annotations and those performed by HUVH cardiologists, which consistently assigned lower quality grades. As harmonization of quality criteria remains an open challenge, EchoQuality provides a standardized and reproducible framework that may fill this gap.

Additional evidence supporting EchoQuality validity was observed. First, in the HUVH-view dataset, the expert selected a clip with the highest available EchoQuality grade in 90.4% of cases, demonstrating strong agreement between automated quality grading and expert best-clip selection in a prospective, independent analysis. Second, study-level EchoQuality grades were significantly associated with quality assessed during clinical practice, supporting the construct of EchoQuality as a study-level indicator. Finally, quality grades were systematically higher for conventional echocardiography than same-day POCUS, suggesting that EchoQuality has truly learned generalizable image quality features.

EchoQuality grades further demonstrated meaningful demographic and temporal associations. Higher BMI was associated with lower quality, consistent with echocardiography technical limitations^9,41^, while the proportion of low-quality studies decreased over time, as expected by the progressive technology improvements. Interestingly, such temporal shifts may progressively redefine what constitutes low image quality, and may support EchoQuality for data drift monitoring in clinical applications.

Overall, high-quality studies resulted in superior AI model performance in 80% of evaluations. Compared with low-quality studies, the median absolute performance differences in high-quality studies were 0.10 for R^2^, 0.14 and 0.12 for sensitivity in dysfunction and disease detections, 0.12 for view identification accuracy, and 0.03 for Dice Score in LV segmentation. This coherent directionality across heterogeneous cohorts, assessments, and models suggests that EchoQuality grade is a consistent determinant of AI model performance. The largest performance gaps, exceeding 20% in sensitivity between high-and low-quality studies for key conditions, occurred in guideline-derived severity classifications, which directly guide clinical management. Overall, these findings highlight the clinical risk of deploying AI-based tools without explicit quality control.

A particularly important observation was that predictions from low-quality videos tended to regress toward median values and most prevalent classes. For severity classification and disease detection, this behavior was characterized by markedly lower sensitivity and slightly higher specificity in low-quality studies, consistent with a shift toward conservative predictions. This behavior suggests that low quality impacts the amount of signal available and raises substantial concerns about the readiness of these models to flag high-risk patients deserving particularly-careful human evaluation. Importantly, a quality-aware aggregation strategy improved performance in 84% of evaluations, suggesting that safety in echocardiographic AI systems may be enhanced by quality-aware selection and weighting.

When considering the present findings, certain study limitations should be considered. First, EchoQuality was developed using exclusively transthoracic 2D cine B-mode acquisitions and was not evaluated in transesophageal echocardiography, color Doppler imaging, and 1D and 3D echocardiography. Second, the definition of image quality was established through a process performed at a single center and may therefore reflect center-specific perception. Multicenter and inclusive processes should be performed to derive a broadly shared definition of echocardiography image quality. Third, analyses could be affected by the inter-center variability in the clinical definition and computation of echocardiographic measurements. Nonetheless, as most analyses were based on within-center comparisons, this is unlikely to have materially influenced the main findings.

In conclusion, EchoQuality is an automated, openly-available framework to quantify image quality in echocardiography videos. Across multiple international cohorts and tasks, higher image quality was consistently associated with superior performance of state-of-the-art AI models, while low image quality led to systematic deterioration and regression toward population-average predictions. Incorporating awareness of image quality into AI workflows may improve robustness and clinical reliability.

## Data Availability

Access to HUVH, PWH, AUMC and FLEMENGHO raw datasets will be granted on approval by the institutional review boards and data sharing committees. Details on how to request access are available from the corresponding author on reasonable request. MIMIC-IV-Echo and CAMUS are available at their respective web pages. EchoQuality is publicly available on our GitHub page https://github.com/CardiovascularImagingVallHebron/EchoQuality.

## Acknowledgments

We would like to thank the cardiologists, sonographers, and researchers who participated in the co-creation sessions, and the personnel of the IT departments of the different hospitals involved for their invaluable work.

## AI declaration statement

The use of natural language processing and generative AI tools has been limited to minor language edits.

## Contributors

PLG, JGO, LDS, AG contributed to study conception, study design, data collection and analysis and drafted the manuscript. MBA, JPBR, NC, AMG, ME, MJB, FF, HM, LRC, SMG, SZ, YQ, FWA, TK, GTT, APWL, XL, KL, LGG, JRP and IFG contributed to data collection and data analysis. All authors critically revised it for important intellectual content.

## Supplementary Material

**Supplemental Figure 1.**
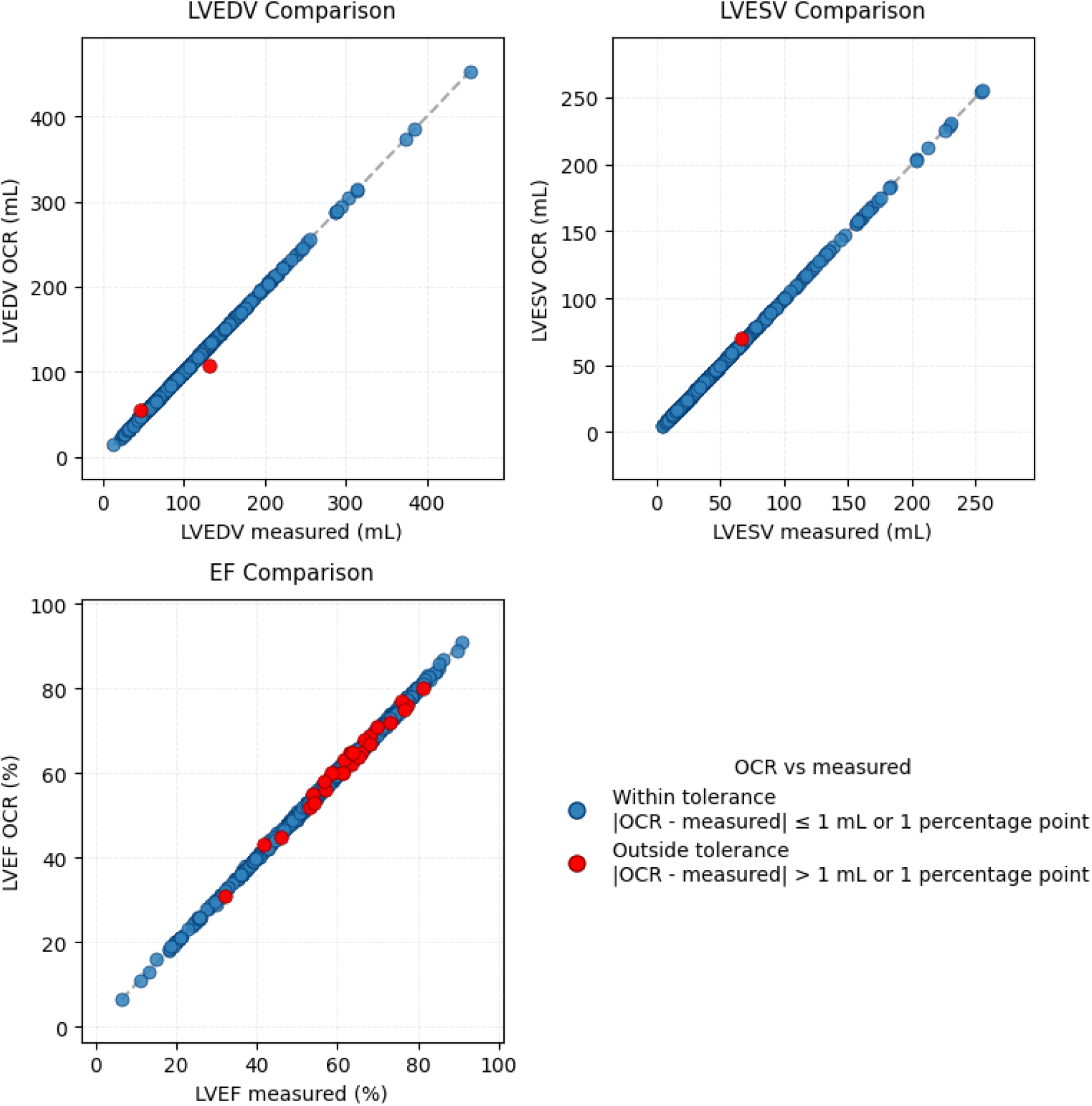
Validation of the optical character recognition (OCR) pipeline used to extract echocardiographic measurements from MIMIC-IV-ECHO images. Agreement between OCR-extracted values and reference labels provided for 1,000 studies in the MIMIC-IV-ECHO-Ext-LVVOLUMES-A4C-ROI ^31,34^ dataset is shown. For LVEDV 2 studies (0.2%) differ by more than 1 mL, for LVESV 1 study (0.1%) differs by more than 1 mL, for LVEF 28 studies (2.8%) differ by more than 1%. classifications. Each row indicates which view types contribute to the prediction of a given measurement or disease.

**Supplemental Table 1.**
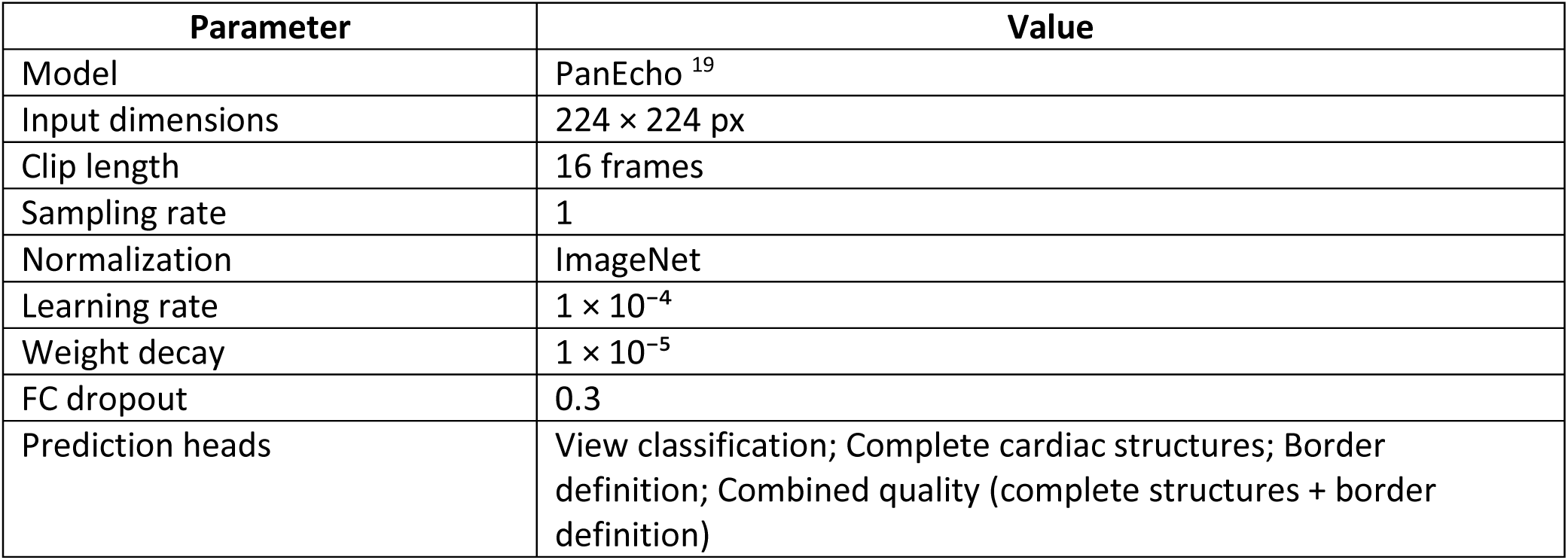
EchoQuality model architecture and training hyperparameters.

| **Parameter** | **Value** |
|---|---|
| Model | PanEcho<sup>19</sup> |
| Input dimensions | 224 × 224 px |
| Clip length | 16 frames |
| Sampling rate | 1 |
| Normalization | ImageNet |
| Learning rate | 1 × 10<sup>−4</sup> |
| Weight decay | 1 × 10<sup>−5</sup> |
| FC dropout | 0.3 |
| Prediction heads | View classification; Complete cardiac structures; Border definition; Combined quality (complete structures + border definition) |

**Supplemental Table 2.** Cohort overview of EchoQuality development and testing.

|  | **Training** | **Internal Testing** | **External Testing** |  |
|---|---|---|---|---|
| Patients, n | 371 | 87 | 500 |  |
| Studies, n | 374 | 89 | 500 |  |
| Videos total, n | 6,937 | 1,903 | 1,000 |  |
| Videos included, n | 5,056 | 1,418 | 1,000 |  |
| Age, median [IQR] | 72 [61-80] | 73 [58-79] | 67 [56-76] |  |
| BMI, median [IQR] | 26.52 [23.74-29.15] | 27.18 [23.02-29.75] | - |  |
| Male sex, n (%) | 203 (55%) | 47 (54%) | 330 (66%) |  |
| LVEF, (%) | 60 [53-64] | 58 [50-63] | 46 [37-52] |  |
| N videos per quality annotated |  |  | **HUVH In-house** | **Native CAMUS** |
| Low, n (%) | 1,617 (32%) | 630 (44%) | 607 (61%) | 116 (12%) |
| Mid, n (%) | 2,750 (54%) | 663 (47%) | 366 (36%) | 379 (38%) |
| High, n (%) | 689 (14%) | 125 (9%) | 27 (3%) | 505 (50%) |

BMI = body mass index, IQR = inter-quartile range, LVEF = left ventricle ejection fraction.

### Clinical-inspired quality-aware aggregation strategy

To incorporate image quality into study-level prediction aggregation, a linear video-level quality weight was computed as (*p_mid + 2 × p_high*) */ 2*, where *p_mid* and *p_high* represent the predicted probabilities of mid and high quality, respectively. This weighting factor was used to balance video-level predictions, such that higher-quality videos contribute proportionally more to the study-level estimates. Videos with a quality score below 0.2 were excluded from aggregation prior to weighting.

**Supplemental Table 3.**
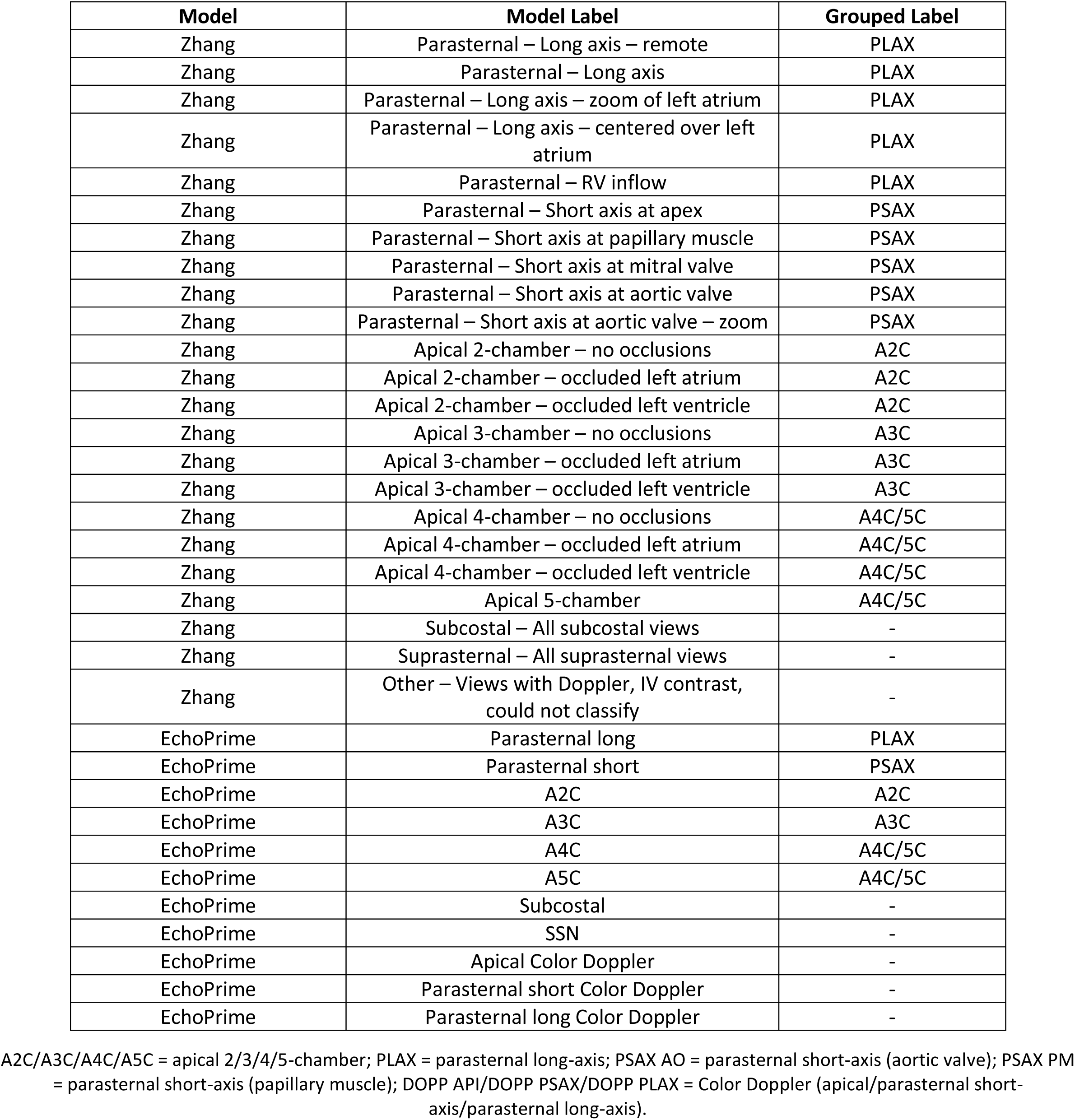
Mapping of view classification labels from the Zhang ^16^ and EchoPrime ^17^ models to the unified comparison labels used in this study.

| Model | Model Label | Grouped Label |
|---|---|---|
| Zhang | Parasternal – Long axis – remote | PLAX |
| Zhang | Parasternal – Long axis | PLAX |
| Zhang | Parasternal – Long axis – zoom of left atrium | PLAX |
| Zhang | Parasternal – Long axis – centered over left atrium | PLAX |
| Zhang | Parasternal – RV inflow | PLAX |
| Zhang | Parasternal – Short axis at apex | PSAX |
| Zhang | Parasternal – Short axis at papillary muscle | PSAX |
| Zhang | Parasternal – Short axis at mitral valve | PSAX |
| Zhang | Parasternal – Short axis at aortic valve | PSAX |
| Zhang | Parasternal – Short axis at aortic valve – zoom | PSAX |
| Zhang | Apical 2-chamber – no occlusions | A2C |
| Zhang | Apical 2-chamber – occluded left atrium | A2C |
| Zhang | Apical 2-chamber – occluded left ventricle | A2C |
| Zhang | Apical 3-chamber – no occlusions | A3C |
| Zhang | Apical 3-chamber – occluded left atrium | A3C |
| Zhang | Apical 3-chamber – occluded left ventricle | A3C |
| Zhang | Apical 4-chamber – no occlusions | A4C/5C |
| Zhang | Apical 4-chamber – occluded left atrium | A4C/5C |
| Zhang | Apical 4-chamber – occluded left ventricle | A4C/5C |
| Zhang | Apical 5-chamber | A4C/5C |
| Zhang | Subcostal – All subcostal views | - |
| Zhang | Suprasternal – All suprasternal views | - |
| Zhang | Other – Views with Doppler, IV contrast, could not classify | - |
| EchoPrime | Parasternal long | PLAX |
| EchoPrime | Parasternal short | PSAX |
| EchoPrime | A2C | A2C |
| EchoPrime | A3C | A3C |
| EchoPrime | A4C | A4C/5C |
| EchoPrime | A5C | A4C/5C |
| EchoPrime | Subcostal | - |
| EchoPrime | SSN | - |
| EchoPrime | Apical Color Doppler | - |
| EchoPrime | Parasternal short Color Doppler | - |
| EchoPrime | Parasternal long Color Doppler | - |

A2C/A3C/A4C/A5C = apical 2/3/4/5-chamber; PLAX = parasternal long-axis; PSAX AO = parasternal short-axis (aortic valve); PSAX PM = parasternal short-axis (papillary muscle); DOPP API/DOPP PSAX/DOPP PLAX = Color Doppler (apical/parasternal short-axis/parasternal long-axis).

**Supplemental Table 4.** Echocardiographic views used by PanEcho for study-level aggregation of measurements and disease.

|  | **A2C** | **A3C** | **A4C** | **A5C** | **PLAX** | **PSAX AO** | **PSAX PM** | **DOPP API** | **DOPP PSAX** | **DOPP PLAX** |
|---|---|---|---|---|---|---|---|---|---|---|
| **Measurements** |  |  |  |  |  |  |  |  |  |  |
| LV Ejection Fraction | **X** | **X** | **X** | **X** | **X** |  | **X** |  |  |  |
| LV End-Diastolic Volume | **X** | **X** | **X** | **X** | **X** |  | **X** |  |  |  |
| LV End-Systolic Volume | **X** | **X** | **X** | **X** | **X** |  | **X** |  |  |  |
| LV Septal Thickness | **X** | **X** | **X** | **X** | **X** |  | **X** |  |  |  |
| LV Posterior Wall Thickness | **X** | **X** | **X** | **X** | **X** |  | **X** |  |  |  |
| LV Diameter (systole) | **X** | **X** | **X** | **X** | **X** |  | **X** |  |  |  |
| LV Diameter (diastole) | **X** | **X** | **X** | **X** | **X** |  | **X** |  |  |  |
| E/e' ratio | **X** | **X** | **X** | **X** | **X** |  |  |  |  |  |
| TAPSE |  |  | **X** | **X** |  | **X** |  |  |  |  |
| LA Diameter (systole) | **X** | **X** | **X** | **X** | **X** |  | **X** |  |  |  |
| LA Volume | **X** | **X** | **X** | **X** | **X** |  | **X** |  |  |  |
| AV Peak Velocity (m/s) |  | **X** |  | **X** | **X** | **X** |  |  |  |  |
| **Disease Classification** |  |  |  |  |  |  |  |  |  |  |
| Pericardial Effusion | **X** | **X** | **X** | **X** | **X** | **X** | **X** |  |  |  |
| Aortic Stenosis |  | **X** |  | **X** | **X** | **X** |  | **X** | **X** | **X** |
| Aortic Regurgitation |  | **X** |  | **X** | **X** | **X** |  | **X** | **X** | **X** |
| Mitral Stenosis | **X** | **X** | **X** | **X** | **X** |  | **X** | **X** | **X** | **X** |
| Mitral Regurgitation | **X** | **X** | **X** | **X** | **X** |  | **X** | **X** | **X** | **X** |
| Tricuspid Regurgitation |  |  | **X** | **X** |  |  |  | **X** |  |  |

A2C/A3C/A4C/A5C = apical 2/3/4/5-chamber; PLAX = parasternal long-axis; PSAX AO = parasternal short-axis (aortic valve); PSAX PM = parasternal short-axis (papillary muscle); DOPP API/DOPP PSAX/DOPP PLAX = Color Doppler (apical/parasternal short-axis/parasternal long-axis).

**Supplemental Figure 2.**
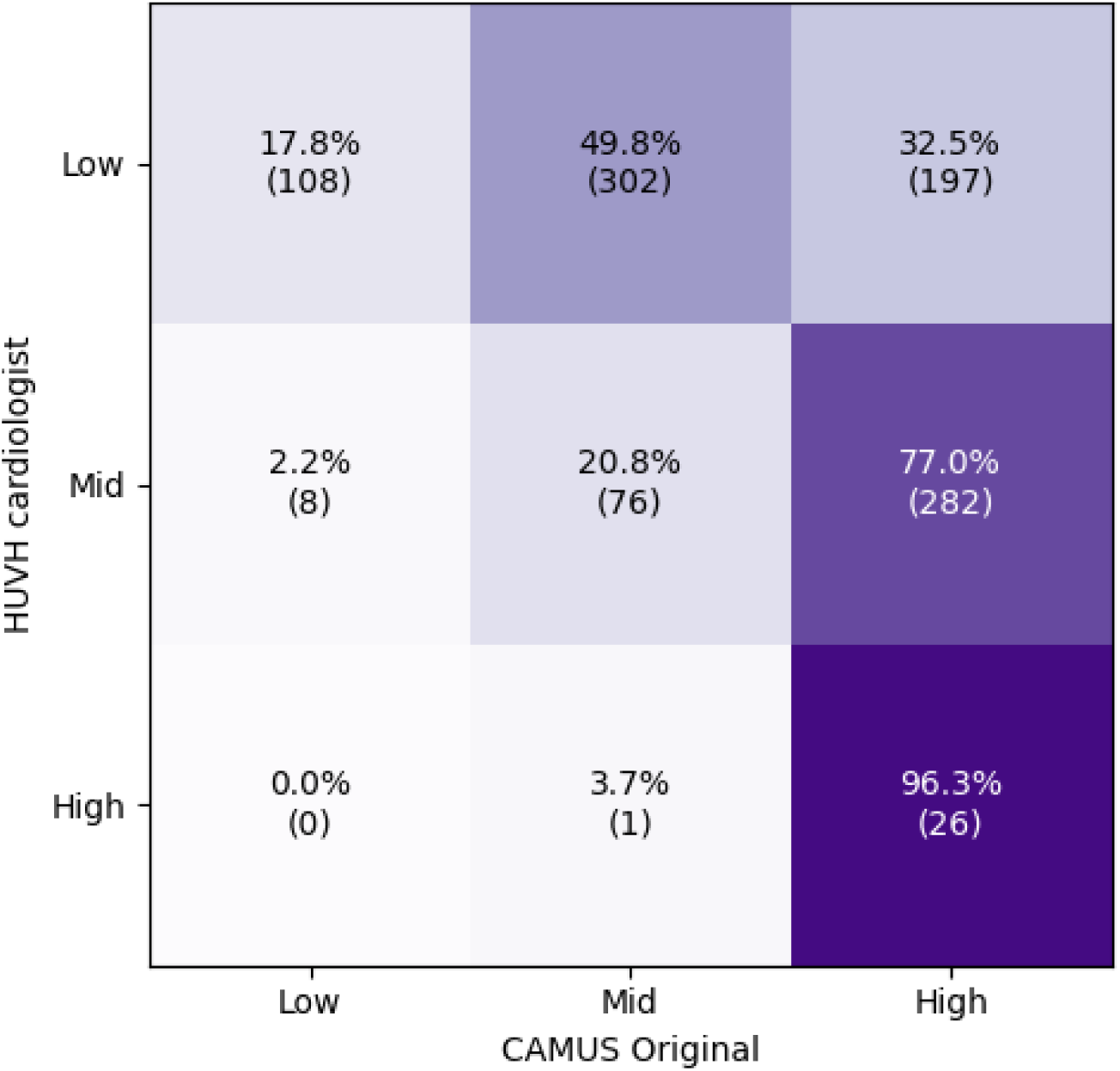
Disagreement between reference standards in the CAMUS external validation dataset.

**Supplemental Figure 3.**
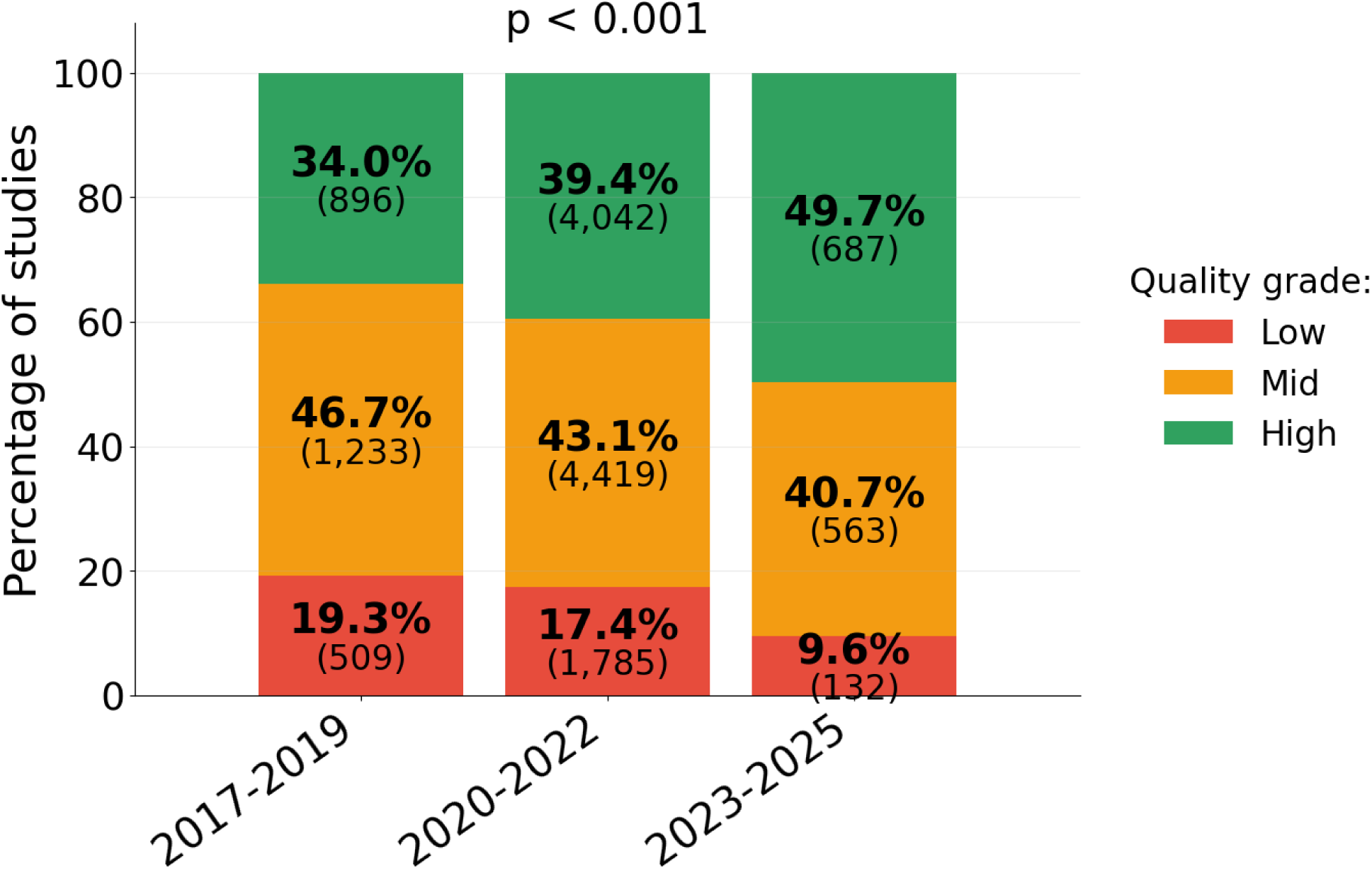
Temporal distribution of EchoQuality grades in the HUVH cohort, showing the proportion of low-, mid-, and high-quality studies across acquisition periods.

**Supplemental Table 5.**
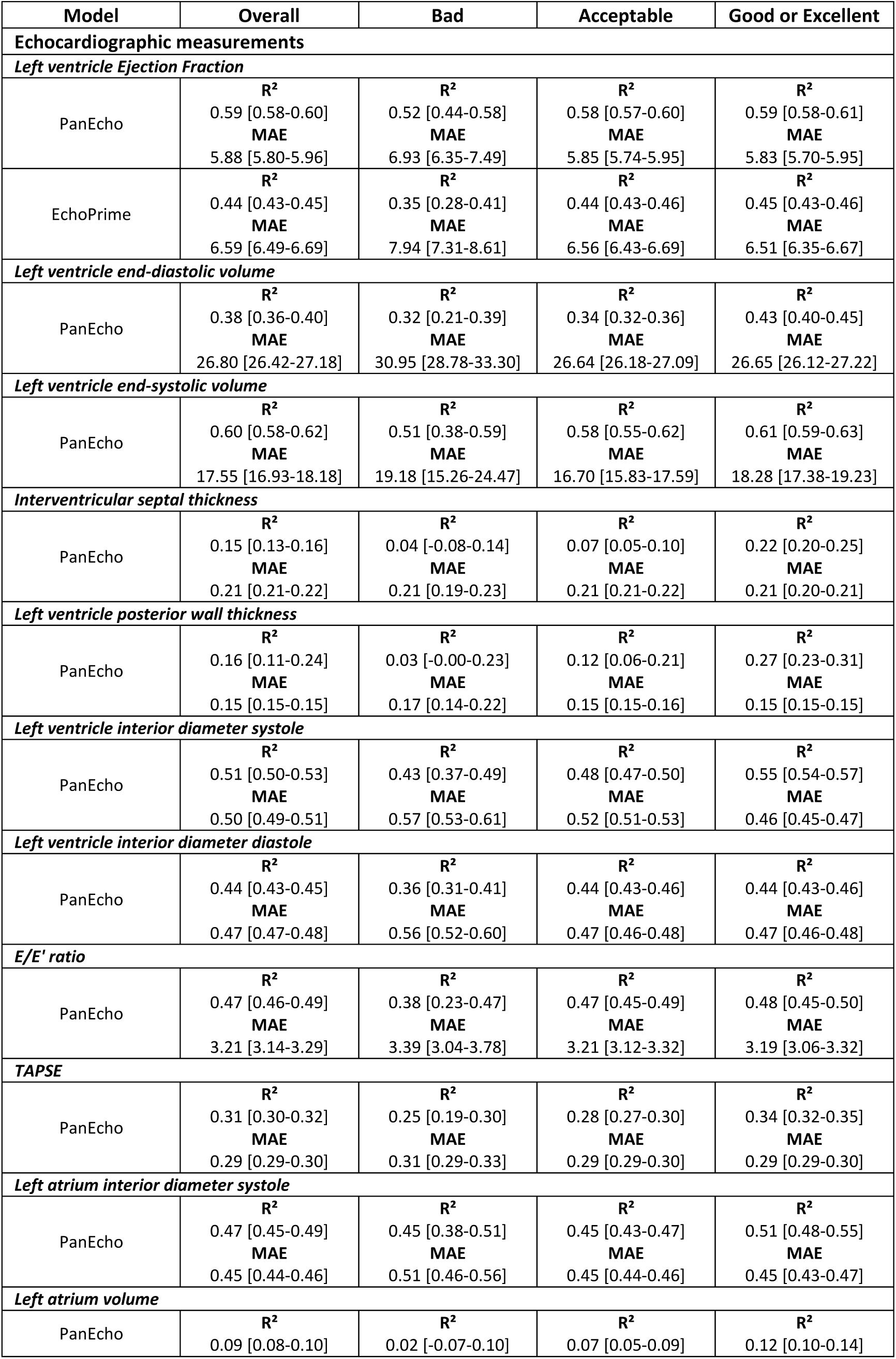
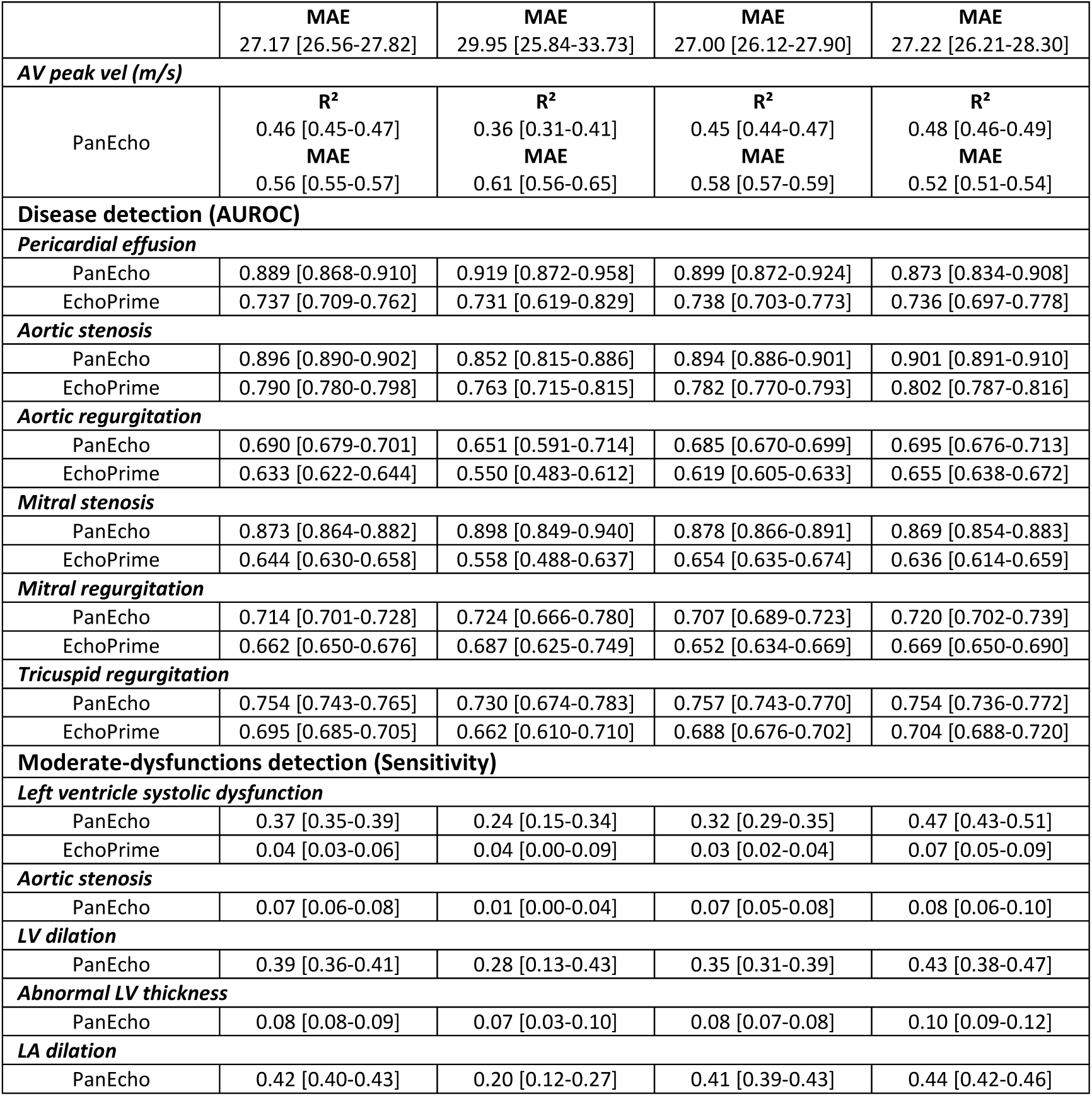
Performance of echocardiographic measurements, moderate dysfunction detection, and disease classification tasks stratified by clinically-annotated quality in HUVH-testing. R², mean absolute error (MAE), area under the receiver operating curve, and sensitivity with 95% confidence intervals are reported for PanEcho and EchoPrime for each measurement, cohort, and clinically-annotated quality grade (bad, acceptable, good or excellent).

| **Model** | **Overall** | **Bad** | **Acceptable** | **Good or Excellent** |
|---|---|---|---|---|
| **Echocardiographic measurements** |  |  |  |  |
| ***Left ventricle Ejection Fraction*** |  |  |  |  |
| PanEcho | **R<sup>2</sup>**<br>0.59 [0.58-0.60]<br>**MAE**<br>5.88 [5.80-5.96] | **R<sup>2</sup>**<br>0.52 [0.44-0.58]<br>**MAE**<br>6.93 [6.35-7.49] | **R<sup>2</sup>**<br>0.58 [0.57-0.60]<br>**MAE**<br>5.85 [5.74-5.95] | **R<sup>2</sup>**<br>0.59 [0.58-0.61]<br>**MAE**<br>5.83 [5.70-5.95] |
| EchoPrime | **R<sup>2</sup>**<br>0.44 [0.43-0.45]<br>**MAE**<br>6.59 [6.49-6.69] | **R<sup>2</sup>**<br>0.35 [0.28-0.41]<br>**MAE**<br>7.94 [7.31-8.61] | **R<sup>2</sup>**<br>0.44 [0.43-0.46]<br>**MAE**<br>6.56 [6.43-6.69] | **R<sup>2</sup>**<br>0.45 [0.43-0.46]<br>**MAE**<br>6.51 [6.35-6.67] |
| ***Left ventricle end-diastolic volume*** |  |  |  |  |
| PanEcho | **R<sup>2</sup>**<br>0.38 [0.36-0.40]<br>**MAE**<br>26.80 [26.42-27.18] | **R<sup>2</sup>**<br>0.32 [0.21-0.39]<br>**MAE**<br>30.95 [28.78-33.30] | **R<sup>2</sup>**<br>0.34 [0.32-0.36]<br>**MAE**<br>26.64 [26.18-27.09] | **R<sup>2</sup>**<br>0.43 [0.40-0.45]<br>**MAE**<br>26.65 [26.12-27.22] |
| ***Left ventricle end-systolic volume*** |  |  |  |  |
| PanEcho | **R<sup>2</sup>**<br>0.60 [0.58-0.62]<br>**MAE**<br>17.55 [16.93-18.18] | **R<sup>2</sup>**<br>0.51 [0.38-0.59]<br>**MAE**<br>19.18 [15.26-24.47] | **R<sup>2</sup>**<br>0.58 [0.55-0.62]<br>**MAE**<br>16.70 [15.83-17.59] | **R<sup>2</sup>**<br>0.61 [0.59-0.63]<br>**MAE**<br>18.28 [17.38-19.23] |
| ***Interventricular septal thickness*** |  |  |  |  |
| PanEcho | **R<sup>2</sup>**<br>0.15 [0.13-0.16]<br>**MAE**<br>0.21 [0.21-0.22] | **R<sup>2</sup>**<br>0.04 [-0.08-0.14]<br>**MAE**<br>0.21 [0.19-0.23] | **R<sup>2</sup>**<br>0.07 [0.05-0.10]<br>**MAE**<br>0.21 [0.21-0.22] | **R<sup>2</sup>**<br>0.22 [0.20-0.25]<br>**MAE**<br>0.21 [0.20-0.21] |
| ***Left ventricle posterior wall thickness*** |  |  |  |  |
| PanEcho | **R<sup>2</sup>**<br>0.16 [0.11-0.24]<br>**MAE**<br>0.15 [0.15-0.15] | **R<sup>2</sup>**<br>0.03 [-0.00-0.23]<br>**MAE**<br>0.17 [0.14-0.22] | **R<sup>2</sup>**<br>0.12 [0.06-0.21]<br>**MAE**<br>0.15 [0.15-0.16] | **R<sup>2</sup>**<br>0.27 [0.23-0.31]<br>**MAE**<br>0.15 [0.15-0.15] |
| ***Left ventricle interior diameter systole*** |  |  |  |  |
| PanEcho | **R<sup>2</sup>**<br>0.51 [0.50-0.53]<br>**MAE**<br>0.50 [0.49-0.51] | **R<sup>2</sup>**<br>0.43 [0.37-0.49]<br>**MAE**<br>0.57 [0.53-0.61] | **R<sup>2</sup>**<br>0.48 [0.47-0.50]<br>**MAE**<br>0.52 [0.51-0.53] | **R<sup>2</sup>**<br>0.55 [0.54-0.57]<br>**MAE**<br>0.46 [0.45-0.47] |
| ***Left ventricle interior diameter diastole*** |  |  |  |  |
| PanEcho | **R<sup>2</sup>**<br>0.44 [0.43-0.45]<br>**MAE**<br>0.47 [0.47-0.48] | **R<sup>2</sup>**<br>0.36 [0.31-0.41]<br>**MAE**<br>0.56 [0.52-0.60] | **R<sup>2</sup>**<br>0.44 [0.43-0.46]<br>**MAE**<br>0.47 [0.46-0.48] | **R<sup>2</sup>**<br>0.44 [0.43-0.46]<br>**MAE**<br>0.47 [0.46-0.48] |
| ***E/E' ratio*** |  |  |  |  |
| PanEcho | **R<sup>2</sup>**<br>0.47 [0.46-0.49]<br>**MAE**<br>3.21 [3.14-3.29] | **R<sup>2</sup>**<br>0.38 [0.23-0.47]<br>**MAE**<br>3.39 [3.04-3.78] | **R<sup>2</sup>**<br>0.47 [0.45-0.49]<br>**MAE**<br>3.21 [3.12-3.32] | **R<sup>2</sup>**<br>0.48 [0.45-0.50]<br>**MAE**<br>3.19 [3.06-3.32] |
| ***TAPSE*** |  |  |  |  |
| PanEcho | **R<sup>2</sup>**<br>0.31 [0.30-0.32]<br>**MAE**<br>0.29 [0.29-0.30] | **R<sup>2</sup>**<br>0.25 [0.19-0.30]<br>**MAE**<br>0.31 [0.29-0.33] | **R<sup>2</sup>**<br>0.28 [0.27-0.30]<br>**MAE**<br>0.29 [0.29-0.30] | **R<sup>2</sup>**<br>0.34 [0.32-0.35]<br>**MAE**<br>0.29 [0.29-0.30] |
| ***Left atrium interior diameter systole*** |  |  |  |  |
| PanEcho | **R<sup>2</sup>**<br>0.47 [0.45-0.49]<br>**MAE**<br>0.45 [0.44-0.46] | **R<sup>2</sup>**<br>0.45 [0.38-0.51]<br>**MAE**<br>0.51 [0.46-0.56] | **R<sup>2</sup>**<br>0.45 [0.43-0.47]<br>**MAE**<br>0.45 [0.44-0.46] | **R<sup>2</sup>**<br>0.51 [0.48-0.55]<br>**MAE**<br>0.45 [0.43-0.47] |
| ***Left atrium volume*** |  |  |  |  |
| PanEcho | **R<sup>2</sup>**<br>0.09 [0.08-0.10]<br>**MAE**<br>27.17 [26.56-27.82] | **R<sup>2</sup>**<br>0.02 [-0.07-0.10]<br>**MAE**<br>29.95 [25.84-33.73] | **R<sup>2</sup>**<br>0.07 [0.05-0.09]<br>**MAE**<br>27.00 [26.12-27.90] | **R<sup>2</sup>**<br>0.12 [0.10-0.14]<br>**MAE**<br>27.22 [26.21-28.30] |
| ***AV peak vel (m/s)*** |  |  |  |  |
| PanEcho | **R<sup>2</sup>**<br>0.46 [0.45-0.47]<br>**MAE**<br>0.56 [0.55-0.57] | **R<sup>2</sup>**<br>0.36 [0.31-0.41]<br>**MAE**<br>0.61 [0.56-0.65] | **R<sup>2</sup>**<br>0.45 [0.44-0.47]<br>**MAE**<br>0.58 [0.57-0.59] | **R<sup>2</sup>**<br>0.48 [0.46-0.49]<br>**MAE**<br>0.52 [0.51-0.54] |
| **Disease detection (AUROC)** |  |  |  |  |
| ***Pericardial effusion*** |  |  |  |  |
| PanEcho | 0.889 [0.868-0.910] | 0.919 [0.872-0.958] | 0.899 [0.872-0.924] | 0.873 [0.834-0.908] |
| EchoPrime | 0.737 [0.709-0.762] | 0.731 [0.619-0.829] | 0.738 [0.703-0.773] | 0.736 [0.697-0.778] |
| ***Aortic stenosis*** |  |  |  |  |
| PanEcho | 0.896 [0.890-0.902] | 0.852 [0.815-0.886] | 0.894 [0.886-0.901] | 0.901 [0.891-0.910] |
| EchoPrime | 0.790 [0.780-0.798] | 0.763 [0.715-0.815] | 0.782 [0.770-0.793] | 0.802 [0.787-0.816] |
| ***Aortic regurgitation*** |  |  |  |  |
| PanEcho | 0.690 [0.679-0.701] | 0.651 [0.591-0.714] | 0.685 [0.670-0.699] | 0.695 [0.676-0.713] |
| EchoPrime | 0.633 [0.622-0.644] | 0.550 [0.483-0.612] | 0.619 [0.605-0.633] | 0.655 [0.638-0.672] |
| ***Mitral stenosis*** |  |  |  |  |
| PanEcho | 0.873 [0.864-0.882] | 0.898 [0.849-0.940] | 0.878 [0.866-0.891] | 0.869 [0.854-0.883] |
| EchoPrime | 0.644 [0.630-0.658] | 0.558 [0.488-0.637] | 0.654 [0.635-0.674] | 0.636 [0.614-0.659] |
| ***Mitral regurgitation*** |  |  |  |  |
| PanEcho | 0.714 [0.701-0.728] | 0.724 [0.666-0.780] | 0.707 [0.689-0.723] | 0.720 [0.702-0.739] |
| EchoPrime | 0.662 [0.650-0.676] | 0.687 [0.625-0.749] | 0.652 [0.634-0.669] | 0.669 [0.650-0.690] |
| ***Tricuspid regurgitation*** |  |  |  |  |
| PanEcho | 0.754 [0.743-0.765] | 0.730 [0.674-0.783] | 0.757 [0.743-0.770] | 0.754 [0.736-0.772] |
| EchoPrime | 0.695 [0.685-0.705] | 0.662 [0.610-0.710] | 0.688 [0.676-0.702] | 0.704 [0.688-0.720] |
| **Moderate-dysfunctions detection (Sensitivity)** |  |  |  |  |
| ***Left ventricle systolic dysfunction*** |  |  |  |  |
| PanEcho | 0.37 [0.35-0.39] | 0.24 [0.15-0.34] | 0.32 [0.29-0.35] | 0.47 [0.43-0.51] |
| EchoPrime | 0.04 [0.03-0.06] | 0.04 [0.00-0.09] | 0.03 [0.02-0.04] | 0.07 [0.05-0.09] |
| ***Aortic stenosis*** |  |  |  |  |
| PanEcho | 0.07 [0.06-0.08] | 0.01 [0.00-0.04] | 0.07 [0.05-0.08] | 0.08 [0.06-0.10] |
| ***LV dilation*** |  |  |  |  |
| PanEcho | 0.39 [0.36-0.41] | 0.28 [0.13-0.43] | 0.35 [0.31-0.39] | 0.43 [0.38-0.47] |
| ***Abnormal LV thickness*** |  |  |  |  |
| PanEcho | 0.08 [0.08-0.09] | 0.07 [0.03-0.10] | 0.08 [0.07-0.08] | 0.10 [0.09-0.12] |
| ***LA dilation*** |  |  |  |  |
| PanEcho | 0.42 [0.40-0.43] | 0.20 [0.12-0.27] | 0.41 [0.39-0.43] | 0.44 [0.42-0.46] |

**Supplemental Figure 4.**
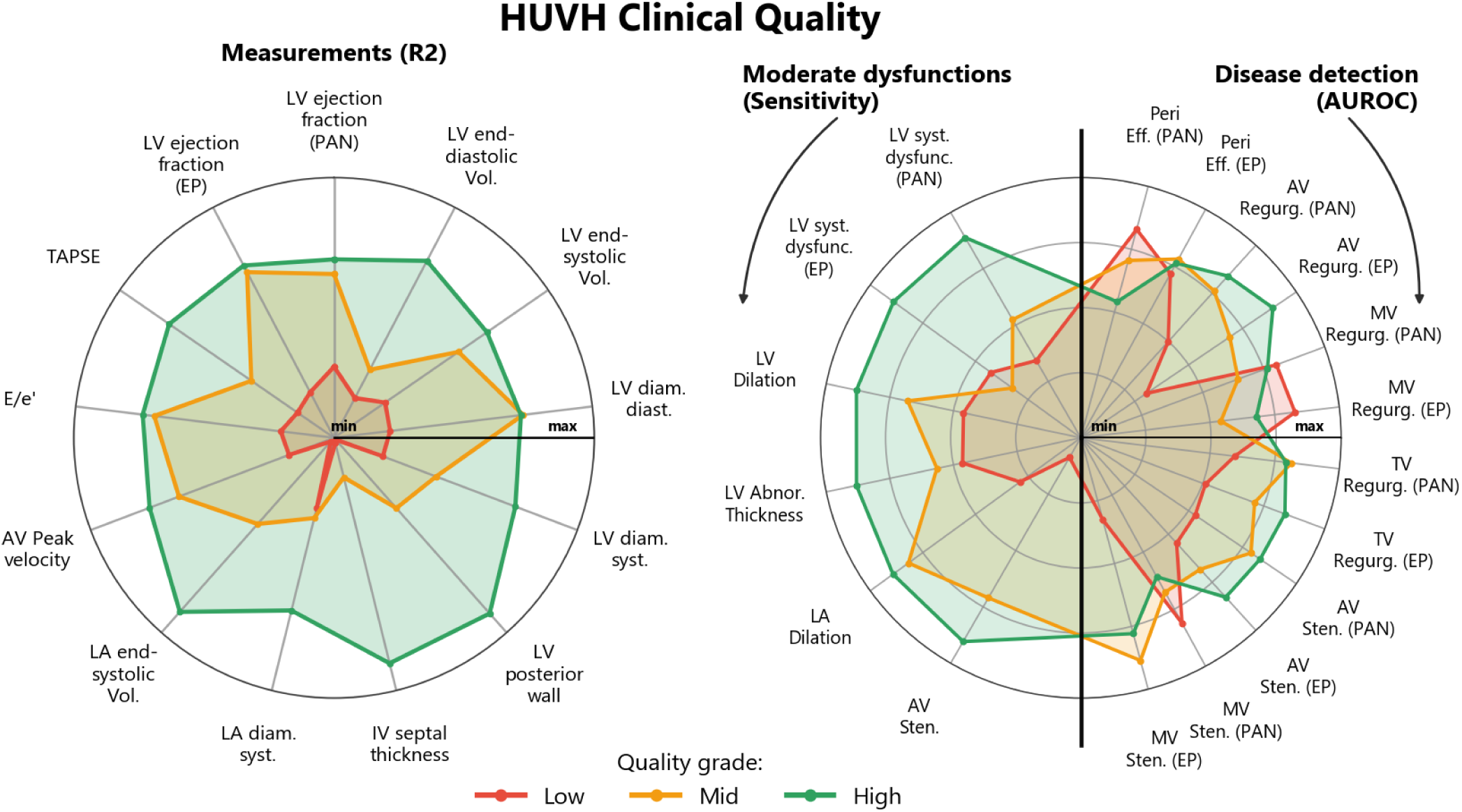
Performance of PanEcho (PAN) and EchoPrime (EP) in echocardiographic measurements, moderate dysfunction detection and disease classification tasks stratified by reported clinical quality in HUVH-testing.

**Supplemental Table 6.**
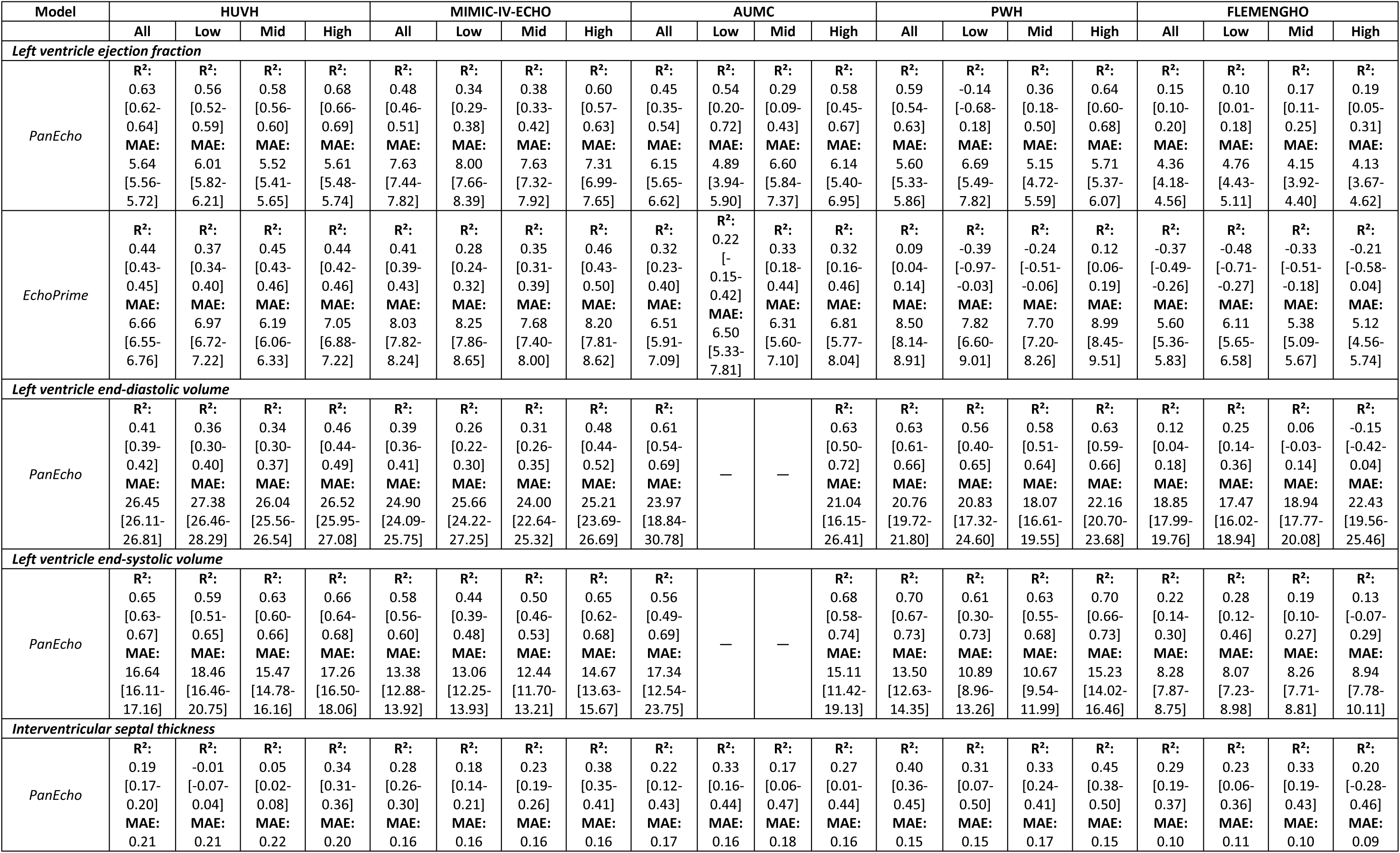
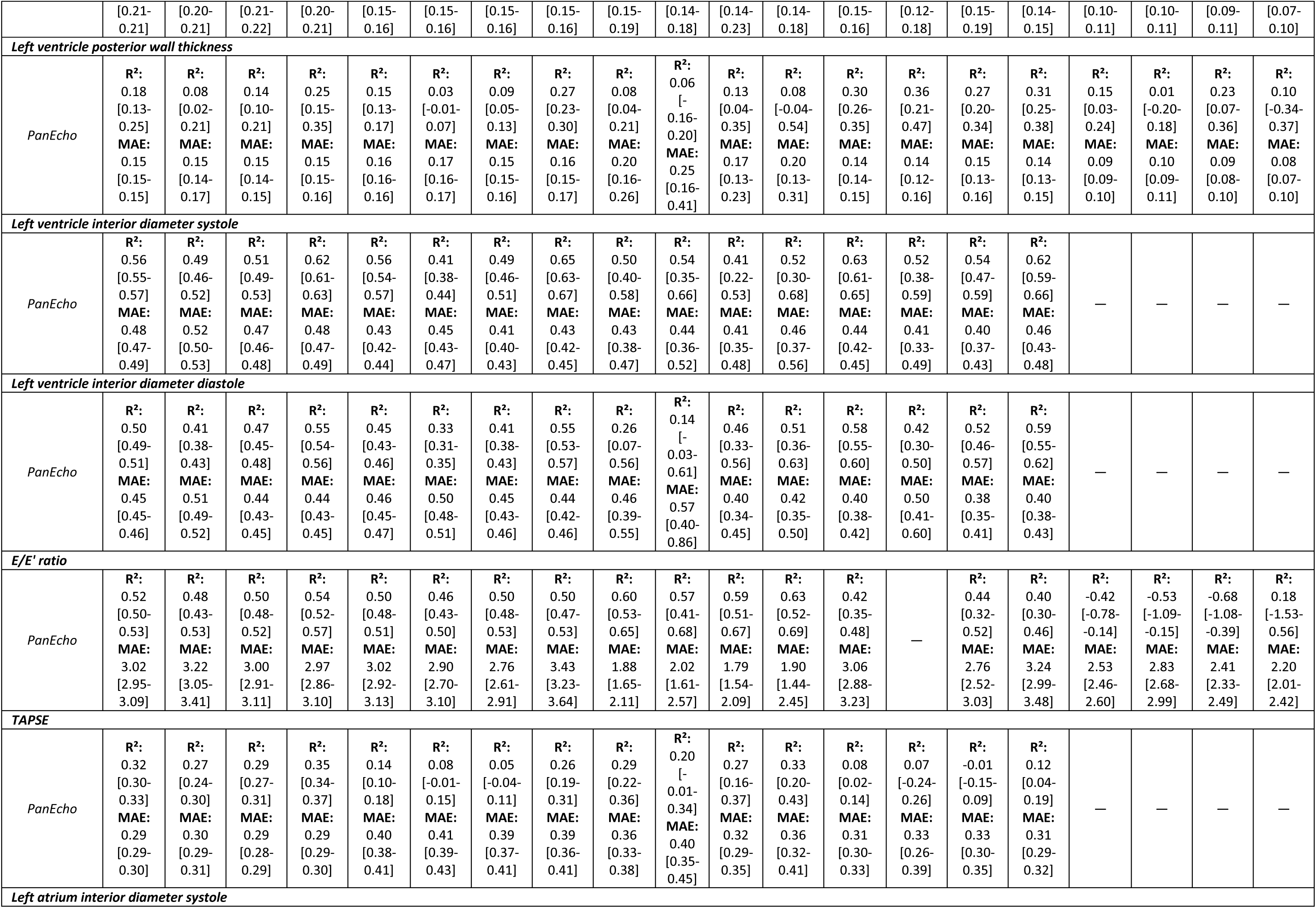
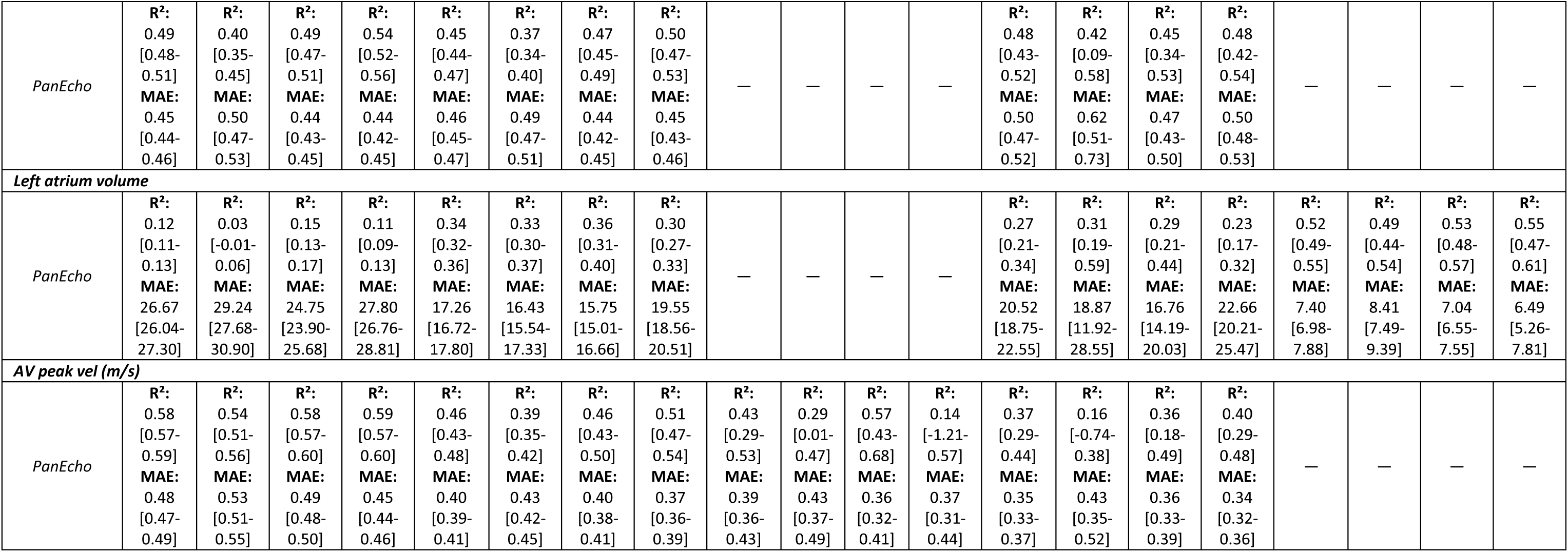
Echocardiographic measurement performance stratified by EchoQuality grade across five cohorts. R² and mean absolute error (MAE) with 95% confidence intervals are reported for PanEcho and EchoPrime for each measurement, cohort, and quality grade (low, mid, high).

| **Model** | **HUVH** |  |  |  | **MIMIC-IV-ECHO** |  |  |  | **AUMC** |  |  |  | **PWH** |  |  |  | **FLEMENGHO** |  |  |  |
|---|---|---|---|---|---|---|---|---|---|---|---|---|---|---|---|---|---|---|---|---|
|  | **All** | **Low** | **Mid** | **High** | **All** | **Low** | **Mid** | **High** | **All** | **Low** | **Mid** | **High** | **All** | **Low** | **Mid** | **High** | **All** | **Low** | **Mid** | **High** |
| ***Left ventricle ejection fraction*** |  |  |  |  |  |  |  |  |  |  |  |  |  |  |  |  |  |  |  |  |
| *PanEcho* | **R<sup>2</sup>:** 0.63 [0.62-0.64]<br>**MAE:** 5.64 [5.56-5.72] | **R<sup>2</sup>:** 0.56 [0.52-0.59]<br>**MAE:** 6.01 [5.82-6.21] | **R<sup>2</sup>:** 0.58 [0.56-0.60]<br>**MAE:** 5.52 [5.41-5.65] | **R<sup>2</sup>:** 0.68 [0.66-0.69]<br>**MAE:** 5.61 [5.48-5.74] | **R<sup>2</sup>:** 0.48 [0.46-0.51]<br>**MAE:** 7.63 [7.44-7.82] | **R<sup>2</sup>:** 0.34 [0.29-0.38]<br>**MAE:** 8.00 [7.66-8.39] | **R<sup>2</sup>:** 0.38 [0.33-0.42]<br>**MAE:** 7.63 [7.32-7.92] | **R<sup>2</sup>:** 0.60 [0.57-0.63]<br>**MAE:** 7.31 [6.99-7.65] | **R<sup>2</sup>:** 0.45 [0.35-0.54]<br>**MAE:** 6.15 [5.65-6.62] | **R<sup>2</sup>:** 0.54 [0.20-0.72]<br>**MAE:** 4.89 [3.94-5.90] | **R<sup>2</sup>:** 0.29 [0.09-0.43]<br>**MAE:** 6.60 [5.84-7.37] | **R<sup>2</sup>:** 0.58 [0.45-0.67]<br>**MAE:** 6.14 [5.40-6.95] | **R<sup>2</sup>:** 0.59 [0.54-0.63]<br>**MAE:** 5.60 [5.33-5.86] | **R<sup>2</sup>:** -0.14 [-0.68-0.18]<br>**MAE:** 6.69 [5.49-7.82] | **R<sup>2</sup>:** 0.36 [0.18-0.50]<br>**MAE:** 5.15 [4.72-5.59] | **R<sup>2</sup>:** 0.64 [0.60-0.68]<br>**MAE:** 5.71 [5.37-6.07] | **R<sup>2</sup>:** 0.15 [0.10-0.20]<br>**MAE:** 4.36 [4.18-4.56] | **R<sup>2</sup>:** 0.10 [0.01-0.18]<br>**MAE:** 4.76 [4.43-5.11] | **R<sup>2</sup>:** 0.17 [0.11-0.25]<br>**MAE:** 4.15 [3.92-4.40] | **R<sup>2</sup>:** 0.19 [0.05-0.31]<br>**MAE:** 4.13 [3.67-4.62] |
| *EchoPrime* | **R<sup>2</sup>:** 0.44 [0.43-0.45]<br>**MAE:** 6.66 [6.55-6.76] | **R<sup>2</sup>:** 0.37 [0.34-0.40]<br>**MAE:** 6.97 [6.72-7.22] | **R<sup>2</sup>:** 0.45 [0.43-0.46]<br>**MAE:** 6.19 [6.06-6.33] | **R<sup>2</sup>:** 0.44 [0.42-0.46]<br>**MAE:** 7.05 [6.88-7.22] | **R<sup>2</sup>:** 0.41 [0.39-0.43]<br>**MAE:** 8.03 [7.82-8.24] | **R<sup>2</sup>:** 0.28 [0.24-0.32]<br>**MAE:** 8.25 [7.86-8.65] | **R<sup>2</sup>:** 0.35 [0.31-0.39]<br>**MAE:** 7.68 [7.40-8.00] | **R<sup>2</sup>:** 0.46 [0.43-0.50]<br>**MAE:** 8.20 [7.81-8.62] | **R<sup>2</sup>:** 0.32 [0.23-0.40]<br>**MAE:** 6.51 [5.91-7.09] | **R<sup>2</sup>:** 0.22 [-0.15-0.42]<br>**MAE:** 6.50 [5.33-7.81] | **R<sup>2</sup>:** 0.33 [0.18-0.44]<br>**MAE:** 6.31 [5.60-7.10] | **R<sup>2</sup>:** 0.32 [0.16-0.46]<br>**MAE:** 6.81 [5.77-8.04] | **R<sup>2</sup>:** 0.09 [0.04-0.14]<br>**MAE:** 8.50 [8.14-8.91] | **R<sup>2</sup>:** -0.39 [-0.97--0.03]<br>**MAE:** 7.82 [6.60-9.01] | **R<sup>2</sup>:** -0.24 [-0.51--0.06]<br>**MAE:** 7.70 [7.20-8.26] | **R<sup>2</sup>:** 0.12 [0.06-0.19]<br>**MAE:** 8.99 [8.45-9.51] | **R<sup>2</sup>:** -0.37 [-0.49--0.26]<br>**MAE:** 5.60 [5.36-5.83] | **R<sup>2</sup>:** -0.48 [-0.71--0.27]<br>**MAE:** 6.11 [5.65-6.58] | **R<sup>2</sup>:** -0.33 [-0.51--0.18]<br>**MAE:** 5.38 [5.09-5.67] | **R<sup>2</sup>:** -0.21 [-0.58-0.04]<br>**MAE:** 5.12 [4.56-5.74] |
| ***Left ventricle end-diastolic volume*** |  |  |  |  |  |  |  |  |  |  |  |  |  |  |  |  |  |  |  |  |
| *PanEcho* | **R<sup>2</sup>:** 0.41 [0.39-0.42]<br>**MAE:** 26.45 [26.11-26.81] | **R<sup>2</sup>:** 0.36 [0.30-0.40]<br>**MAE:** 27.38 [26.46-28.29] | **R<sup>2</sup>:** 0.34 [0.30-0.37]<br>**MAE:** 26.04 [25.56-26.54] | **R<sup>2</sup>:** 0.46 [0.44-0.49]<br>**MAE:** 26.52 [25.95-27.08] | **R<sup>2</sup>:** 0.39 [0.36-0.41]<br>**MAE:** 24.90 [24.09-25.75] | **R<sup>2</sup>:** 0.26 [0.22-0.30]<br>**MAE:** 25.66 [24.22-27.25] | **R<sup>2</sup>:** 0.31 [0.26-0.35]<br>**MAE:** 24.00 [22.64-25.32] | **R<sup>2</sup>:** 0.48 [0.44-0.52]<br>**MAE:** 25.21 [23.69-26.69] | **R<sup>2</sup>:** 0.61 [0.54-0.69]<br>**MAE:** 23.97 [18.84-30.78] | — | — | **R<sup>2</sup>:** 0.63 [0.50-0.72]<br>**MAE:** 21.04 [16.15-26.41] | **R<sup>2</sup>:** 0.63 [0.61-0.66]<br>**MAE:** 20.76 [19.72-21.80] | **R<sup>2</sup>:** 0.56 [0.40-0.65]<br>**MAE:** 20.83 [17.32-24.60] | **R<sup>2</sup>:** 0.58 [0.51-0.64]<br>**MAE:** 18.07 [16.61-19.55] | **R<sup>2</sup>:** 0.63 [0.59-0.66]<br>**MAE:** 22.16 [20.70-23.68] | **R<sup>2</sup>:** 0.12 [0.04-0.18]<br>**MAE:** 18.85 [17.99-19.76] | **R<sup>2</sup>:** 0.25 [0.14-0.36]<br>**MAE:** 17.47 [16.02-18.94] | **R<sup>2</sup>:** 0.06 [-0.03-0.14]<br>**MAE:** 18.94 [17.77-20.08] | **R<sup>2</sup>:** -0.15 [-0.42-0.04]<br>**MAE:** 22.43 [19.56-25.46] |
| ***Left ventricle end-systolic volume*** |  |  |  |  |  |  |  |  |  |  |  |  |  |  |  |  |  |  |  |  |
| *PanEcho* | **R<sup>2</sup>:** 0.65 [0.63-0.67]<br>**MAE:** 16.64 [16.11-17.16] | **R<sup>2</sup>:** 0.59 [0.51-0.65]<br>**MAE:** 18.46 [16.46-20.75] | **R<sup>2</sup>:** 0.63 [0.60-0.66]<br>**MAE:** 15.47 [14.78-16.16] | **R<sup>2</sup>:** 0.66 [0.64-0.68]<br>**MAE:** 17.26 [16.50-18.06] | **R<sup>2</sup>:** 0.58 [0.56-0.60]<br>**MAE:** 13.38 [12.88-13.92] | **R<sup>2</sup>:** 0.44 [0.39-0.48]<br>**MAE:** 13.06 [12.25-13.93] | **R<sup>2</sup>:** 0.50 [0.46-0.53]<br>**MAE:** 12.44 [11.70-13.21] | **R<sup>2</sup>:** 0.65 [0.62-0.68]<br>**MAE:** 14.67 [13.63-15.67] | **R<sup>2</sup>:** 0.56 [0.49-0.69]<br>**MAE:** 17.34 [12.54-23.75] | — | — | **R<sup>2</sup>:** 0.68 [0.58-0.74]<br>**MAE:** 15.11 [11.42-19.13] | **R<sup>2</sup>:** 0.70 [0.67-0.73]<br>**MAE:** 13.50 [12.63-14.35] | **R<sup>2</sup>:** 0.61 [0.30-0.73]<br>**MAE:** 10.89 [8.96-13.26] | **R<sup>2</sup>:** 0.63 [0.55-0.68]<br>**MAE:** 10.67 [9.54-11.99] | **R<sup>2</sup>:** 0.70 [0.66-0.73]<br>**MAE:** 15.23 [14.02-16.46] | **R<sup>2</sup>:** 0.22 [0.14-0.30]<br>**MAE:** 8.28 [7.87-8.75] | **R<sup>2</sup>:** 0.28 [0.12-0.46]<br>**MAE:** 8.07 [7.23-8.98] | **R<sup>2</sup>:** 0.19 [0.10-0.27]<br>**MAE:** 8.26 [7.71-8.81] | **R<sup>2</sup>:** 0.13 [-0.07-0.29]<br>**MAE:** 8.94 [7.78-10.11] |
| ***Interventricular septal thickness*** |  |  |  |  |  |  |  |  |  |  |  |  |  |  |  |  |  |  |  |  |
| *PanEcho* | **R<sup>2</sup>:** 0.19 [0.17-0.20]<br>**MAE:** 0.21 [0.21-0.21] | **R<sup>2</sup>:** -0.01 [-0.07-0.04]<br>**MAE:** 0.21 [0.20-0.21] | **R<sup>2</sup>:** 0.05 [0.02-0.08]<br>**MAE:** 0.22 [0.21-0.22] | **R<sup>2</sup>:** 0.34 [0.31-0.36]<br>**MAE:** 0.20 [0.20-0.21] | **R<sup>2</sup>:** 0.28 [0.26-0.30]<br>**MAE:** 0.16 [0.15-0.16] | **R<sup>2</sup>:** 0.18 [0.14-0.21]<br>**MAE:** 0.16 [0.15-0.16] | **R<sup>2</sup>:** 0.23 [0.19-0.26]<br>**MAE:** 0.16 [0.15-0.16] | **R<sup>2</sup>:** 0.38 [0.35-0.41]<br>**MAE:** 0.16 [0.15-0.16] | **R<sup>2</sup>:** 0.22 [0.12-0.43]<br>**MAE:** 0.17 [0.15-0.19] | **R<sup>2</sup>:** 0.33 [0.16-0.44]<br>**MAE:** 0.16 [0.14-0.18] | **R<sup>2</sup>:** 0.17 [0.06-0.47]<br>**MAE:** 0.18 [0.14-0.23] | **R<sup>2</sup>:** 0.27 [0.01-0.44]<br>**MAE:** 0.16 [0.14-0.18] | **R<sup>2</sup>:** 0.40 [0.36-0.45]<br>**MAE:** 0.15 [0.15-0.16] | **R<sup>2</sup>:** 0.31 [0.07-0.50]<br>**MAE:** 0.15 [0.12-0.18] | **R<sup>2</sup>:** 0.33 [0.24-0.41]<br>**MAE:** 0.17 [0.15-0.19] | **R<sup>2</sup>:** 0.45 [0.38-0.50]<br>**MAE:** 0.15 [0.14-0.15] | **R<sup>2</sup>:** 0.29 [0.19-0.37]<br>**MAE:** 0.10 [0.10-0.11] | **R<sup>2</sup>:** 0.23 [0.06-0.36]<br>**MAE:** 0.11 [0.10-0.11] | **R<sup>2</sup>:** 0.33 [0.19-0.43]<br>**MAE:** 0.10 [0.09-0.11] | **R<sup>2</sup>:** 0.20 [-0.28-0.46]<br>**MAE:** 0.09 [0.07-0.10] |
| ***Left ventricle posterior wall thickness*** |  |  |  |  |  |  |  |  |  |  |  |  |  |  |  |  |  |  |  |  |
| *PanEcho* | **R<sup>2</sup>:** 0.18 [0.13-0.25]<br>**MAE:** 0.15 [0.15-0.15] | **R<sup>2</sup>:** 0.08 [0.02-0.21]<br>**MAE:** 0.15 [0.14-0.17] | **R<sup>2</sup>:** 0.14 [0.10-0.21]<br>**MAE:** 0.15 [0.14-0.15] | **R<sup>2</sup>:** 0.25 [0.15-0.35]<br>**MAE:** 0.15 [0.15-0.16] | **R<sup>2</sup>:** 0.15 [0.13-0.17]<br>**MAE:** 0.16 [0.16-0.16] | **R<sup>2</sup>:** 0.03 [-0.01-0.07]<br>**MAE:** 0.17 [0.16-0.17] | **R<sup>2</sup>:** 0.09 [0.05-0.13]<br>**MAE:** 0.15 [0.15-0.16] | **R<sup>2</sup>:** 0.27 [0.23-0.30]<br>**MAE:** 0.16 [0.15-0.17] | **R<sup>2</sup>:** 0.08 [0.04-0.21]<br>**MAE:** 0.20 [0.16-0.26] | **R<sup>2</sup>:** 0.06 [-0.16-0.20]<br>**MAE:** 0.25 [0.16-0.41] | **R<sup>2</sup>:** 0.13 [0.04-0.35]<br>**MAE:** 0.17 [0.13-0.23] | **R<sup>2</sup>:** 0.08 [-0.04-0.54]<br>**MAE:** 0.20 [0.13-0.31] | **R<sup>2</sup>:** 0.30 [0.26-0.35]<br>**MAE:** 0.14 [0.14-0.15] | **R<sup>2</sup>:** 0.36 [0.21-0.47]<br>**MAE:** 0.14 [0.12-0.16] | **R<sup>2</sup>:** 0.27 [0.20-0.34]<br>**MAE:** 0.15 [0.13-0.16] | **R<sup>2</sup>:** 0.31 [0.25-0.38]<br>**MAE:** 0.14 [0.13-0.15] | **R<sup>2</sup>:** 0.15 [0.03-0.24]<br>**MAE:** 0.09 [0.09-0.10] | **R<sup>2</sup>:** 0.01 [-0.20-0.18]<br>**MAE:** 0.10 [0.09-0.11] | **R<sup>2</sup>:** 0.23 [0.07-0.36]<br>**MAE:** 0.09 [0.08-0.10] | **R<sup>2</sup>:** 0.10 [-0.34-0.37]<br>**MAE:** 0.08 [0.07-0.10] |
| ***Left ventricle interior diameter systole*** |  |  |  |  |  |  |  |  |  |  |  |  |  |  |  |  |  |  |  |  |
| *PanEcho* | **R<sup>2</sup>:** 0.56 [0.55-0.57]<br>**MAE:** 0.48 [0.47-0.49] | **R<sup>2</sup>:** 0.49 [0.46-0.52]<br>**MAE:** 0.52 [0.50-0.53] | **R<sup>2</sup>:** 0.51 [0.49-0.53]<br>**MAE:** 0.47 [0.46-0.48] | **R<sup>2</sup>:** 0.62 [0.61-0.63]<br>**MAE:** 0.48 [0.47-0.49] | **R<sup>2</sup>:** 0.56 [0.54-0.57]<br>**MAE:** 0.43 [0.42-0.44] | **R<sup>2</sup>:** 0.41 [0.38-0.44]<br>**MAE:** 0.45 [0.43-0.47] | **R<sup>2</sup>:** 0.49 [0.46-0.51]<br>**MAE:** 0.41 [0.40-0.43] | **R<sup>2</sup>:** 0.65 [0.63-0.67]<br>**MAE:** 0.43 [0.42-0.45] | **R<sup>2</sup>:** 0.50 [0.40-0.58]<br>**MAE:** 0.43 [0.38-0.47] | **R<sup>2</sup>:** 0.54 [0.35-0.66]<br>**MAE:** 0.44 [0.36-0.52] | **R<sup>2</sup>:** 0.41 [0.22-0.53]<br>**MAE:** 0.41 [0.35-0.48] | **R<sup>2</sup>:** 0.52 [0.30-0.68]<br>**MAE:** 0.46 [0.37-0.56] | **R<sup>2</sup>:** 0.63 [0.61-0.65]<br>**MAE:** 0.44 [0.42-0.45] | **R<sup>2</sup>:** 0.52 [0.38-0.59]<br>**MAE:** 0.41 [0.33-0.49] | **R<sup>2</sup>:** 0.54 [0.47-0.59]<br>**MAE:** 0.40 [0.37-0.43] | **R<sup>2</sup>:** 0.62 [0.59-0.66]<br>**MAE:** 0.46 [0.43-0.48] | — | — | — | — |
| ***Left ventricle interior diameter diastole*** |  |  |  |  |  |  |  |  |  |  |  |  |  |  |  |  |  |  |  |  |
| *PanEcho* | **R<sup>2</sup>:** 0.50 [0.49-0.51]<br>**MAE:** 0.45 [0.45-0.46] | **R<sup>2</sup>:** 0.41 [0.38-0.43]<br>**MAE:** 0.51 [0.49-0.52] | **R<sup>2</sup>:** 0.47 [0.45-0.48]<br>**MAE:** 0.44 [0.43-0.45] | **R<sup>2</sup>:** 0.55 [0.54-0.56]<br>**MAE:** 0.44 [0.43-0.45] | **R<sup>2</sup>:** 0.45 [0.43-0.46]<br>**MAE:** 0.46 [0.45-0.47] | **R<sup>2</sup>:** 0.33 [0.31-0.35]<br>**MAE:** 0.50 [0.48-0.51] | **R<sup>2</sup>:** 0.41 [0.38-0.43]<br>**MAE:** 0.45 [0.43-0.46] | **R<sup>2</sup>:** 0.55 [0.53-0.57]<br>**MAE:** 0.44 [0.42-0.46] | **R<sup>2</sup>:** 0.26 [0.07-0.56]<br>**MAE:** 0.46 [0.39-0.55] | **R<sup>2</sup>:** 0.14 [-0.03-0.61]<br>**MAE:** 0.57 [0.40-0.86] | **R<sup>2</sup>:** 0.46 [0.33-0.56]<br>**MAE:** 0.40 [0.34-0.45] | **R<sup>2</sup>:** 0.51 [0.36-0.63]<br>**MAE:** 0.42 [0.35-0.50] | **R<sup>2</sup>:** 0.58 [0.55-0.60]<br>**MAE:** 0.40 [0.38-0.42] | **R<sup>2</sup>:** 0.42 [0.30-0.50]<br>**MAE:** 0.50 [0.41-0.60] | **R<sup>2</sup>:** 0.52 [0.46-0.57]<br>**MAE:** 0.38 [0.35-0.41] | **R<sup>2</sup>:** 0.59 [0.55-0.62]<br>**MAE:** 0.40 [0.38-0.43] | — | — | — | — |
| ***E/E' ratio*** |  |  |  |  |  |  |  |  |  |  |  |  |  |  |  |  |  |  |  |  |
| *PanEcho* | **R<sup>2</sup>:** 0.52 [0.50-0.53]<br>**MAE:** 3.02 [2.95-3.09] | **R<sup>2</sup>:** 0.48 [0.43-0.53]<br>**MAE:** 3.22 [3.05-3.41] | **R<sup>2</sup>:** 0.50 [0.48-0.52]<br>**MAE:** 3.00 [2.91-3.11] | **R<sup>2</sup>:** 0.54 [0.52-0.57]<br>**MAE:** 2.97 [2.86-3.10] | **R<sup>2</sup>:** 0.50 [0.48-0.51]<br>**MAE:** 3.02 [2.92-3.13] | **R<sup>2</sup>:** 0.46 [0.43-0.50]<br>**MAE:** 2.90 [2.70-3.10] | **R<sup>2</sup>:** 0.50 [0.48-0.53]<br>**MAE:** 2.76 [2.61-2.91] | **R<sup>2</sup>:** 0.50 [0.47-0.53]<br>**MAE:** 3.43 [3.23-3.64] | **R<sup>2</sup>:** 0.60 [0.53-0.65]<br>**MAE:** 1.88 [1.65-2.11] | **R<sup>2</sup>:** 0.57 [0.41-0.68]<br>**MAE:** 2.02 [1.61-2.57] | **R<sup>2</sup>:** 0.59 [0.51-0.67]<br>**MAE:** 1.79 [1.54-2.09] | **R<sup>2</sup>:** 0.63 [0.52-0.69]<br>**MAE:** 1.90 [1.44-2.45] | **R<sup>2</sup>:** 0.42 [0.35-0.48]<br>**MAE:** 3.06 [2.88-3.23] | — | **R<sup>2</sup>:** 0.44 [0.32-0.52]<br>**MAE:** 2.76 [2.52-3.03] | **R<sup>2</sup>:** 0.40 [0.30-0.46]<br>**MAE:** 3.24 [2.99-3.48] | **R<sup>2</sup>:** -0.42 [-0.78--0.14]<br>**MAE:** 2.53 [2.46-2.60] | **R<sup>2</sup>:** -0.53 [-1.09--0.15]<br>**MAE:** 2.83 [2.68-2.99] | **R<sup>2</sup>:** -0.68 [-1.08--0.39]<br>**MAE:** 2.41 [2.33-2.49] | **R<sup>2</sup>:** 0.18 [-1.53-0.56]<br>**MAE:** 2.20 [2.01-2.42] |
| ***TAPSE*** |  |  |  |  |  |  |  |  |  |  |  |  |  |  |  |  |  |  |  |  |
| *PanEcho* | **R<sup>2</sup>:** 0.32 [0.30-0.33]<br>**MAE:** 0.29 [0.29-0.30] | **R<sup>2</sup>:** 0.27 [0.24-0.30]<br>**MAE:** 0.30 [0.29-0.31] | **R<sup>2</sup>:** 0.29 [0.27-0.31]<br>**MAE:** 0.29 [0.28-0.29] | **R<sup>2</sup>:** 0.35 [0.34-0.37]<br>**MAE:** 0.29 [0.29-0.30] | **R<sup>2</sup>:** 0.14 [0.10-0.18]<br>**MAE:** 0.40 [0.38-0.41] | **R<sup>2</sup>:** 0.08 [-0.01-0.15]<br>**MAE:** 0.41 [0.39-0.43] | **R<sup>2</sup>:** 0.05 [-0.04-0.11]<br>**MAE:** 0.39 [0.37-0.41] | **R<sup>2</sup>:** 0.26 [0.19-0.31]<br>**MAE:** 0.39 [0.36-0.41] | **R<sup>2</sup>:** 0.29 [0.22-0.36]<br>**MAE:** 0.36 [0.33-0.38] | **R<sup>2</sup>:** 0.20 [-0.01-0.34]<br>**MAE:** 0.40 [0.35-0.45] | **R<sup>2</sup>:** 0.27 [0.16-0.37]<br>**MAE:** 0.32 [0.29-0.35] | **R<sup>2</sup>:** 0.33 [0.20-0.43]<br>**MAE:** 0.36 [0.32-0.41] | **R<sup>2</sup>:** 0.08 [0.02-0.14]<br>**MAE:** 0.31 [0.30-0.33] | **R<sup>2</sup>:** 0.07 [-0.24-0.26]<br>**MAE:** 0.33 [0.26-0.39] | **R<sup>2</sup>:** -0.01 [-0.15-0.09]<br>**MAE:** 0.33 [0.30-0.35] | **R<sup>2</sup>:** 0.12 [0.04-0.19]<br>**MAE:** 0.31 [0.29-0.32] | — | — | — | — |
| ***Left atrium interior diameter systole*** |  |  |  |  |  |  |  |  |  |  |  |  |  |  |  |  |  |  |  |  |
| *PanEcho* | **R<sup>2</sup>:** 0.49 [0.48-0.51]<br>**MAE:** 0.45 [0.44-0.46] | **R<sup>2</sup>:** 0.40 [0.35-0.45]<br>**MAE:** 0.50 [0.47-0.53] | **R<sup>2</sup>:** 0.49 [0.47-0.51]<br>**MAE:** 0.44 [0.43-0.45] | **R<sup>2</sup>:** 0.54 [0.52-0.56]<br>**MAE:** 0.44 [0.42-0.45] | **R<sup>2</sup>:** 0.45 [0.44-0.47]<br>**MAE:** 0.46 [0.45-0.47] | **R<sup>2</sup>:** 0.37 [0.34-0.40]<br>**MAE:** 0.49 [0.47-0.51] | **R<sup>2</sup>:** 0.47 [0.45-0.49]<br>**MAE:** 0.44 [0.42-0.45] | **R<sup>2</sup>:** 0.50 [0.47-0.53]<br>**MAE:** 0.45 [0.43-0.46] | — | — | — | — | **R<sup>2</sup>:** 0.48 [0.43-0.52]<br>**MAE:** 0.50 [0.47-0.52] | **R<sup>2</sup>:** 0.42 [0.09-0.58]<br>**MAE:** 0.62 [0.51-0.73] | **R<sup>2</sup>:** 0.45 [0.34-0.53]<br>**MAE:** 0.47 [0.43-0.50] | **R<sup>2</sup>:** 0.48 [0.42-0.54]<br>**MAE:** 0.50 [0.48-0.53] | — | — | — | — |
| ***Left atrium volume*** |  |  |  |  |  |  |  |  |  |  |  |  |  |  |  |  |  |  |  |  |
| *PanEcho* | **R<sup>2</sup>:** 0.12 [0.11-0.13]<br>**MAE:** 26.67 [26.04-27.30] | **R<sup>2</sup>:** 0.03 [-0.01-0.06]<br>**MAE:** 29.24 [27.68-30.90] | **R<sup>2</sup>:** 0.15 [0.13-0.17]<br>**MAE:** 24.75 [23.90-25.68] | **R<sup>2</sup>:** 0.11 [0.09-0.13]<br>**MAE:** 27.80 [26.76-28.81] | **R<sup>2</sup>:** 0.34 [0.32-0.36]<br>**MAE:** 17.26 [16.72-17.80] | **R<sup>2</sup>:** 0.33 [0.30-0.37]<br>**MAE:** 16.43 [15.54-17.33] | **R<sup>2</sup>:** 0.36 [0.31-0.40]<br>**MAE:** 15.75 [15.01-16.66] | **R<sup>2</sup>:** 0.30 [0.27-0.33]<br>**MAE:** 19.55 [18.56-20.51] | — | — | — | — | **R<sup>2</sup>:** 0.27 [0.21-0.34]<br>**MAE:** 20.52 [18.75-22.55] | **R<sup>2</sup>:** 0.31 [0.19-0.59]<br>**MAE:** 18.87 [11.92-28.55] | **R<sup>2</sup>:** 0.29 [0.21-0.44]<br>**MAE:** 16.76 [14.19-20.03] | **R<sup>2</sup>:** 0.23 [0.17-0.32]<br>**MAE:** 22.66 [20.21-25.47] | **R<sup>2</sup>:** 0.52 [0.49-0.55]<br>**MAE:** 7.40 [6.98-7.88] | **R<sup>2</sup>:** 0.49 [0.44-0.54]<br>**MAE:** 8.41 [7.49-9.39] | **R<sup>2</sup>:** 0.53 [0.48-0.57]<br>**MAE:** 7.04 [6.55-7.55] | **R<sup>2</sup>:** 0.55 [0.47-0.61]<br>**MAE:** 6.49 [5.26-7.81] |
| ***AV peak vel (m/s)*** |  |  |  |  |  |  |  |  |  |  |  |  |  |  |  |  |  |  |  |  |
| *PanEcho* | **R<sup>2</sup>:** 0.58 [0.57-0.59]<br>**MAE:** 0.48 [0.47-0.49] | **R<sup>2</sup>:** 0.54 [0.51-0.56]<br>**MAE:** 0.53 [0.51-0.55] | **R<sup>2</sup>:** 0.58 [0.57-0.60]<br>**MAE:** 0.49 [0.48-0.50] | **R<sup>2</sup>:** 0.59 [0.57-0.60]<br>**MAE:** 0.45 [0.44-0.46] | **R<sup>2</sup>:** 0.46 [0.43-0.48]<br>**MAE:** 0.40 [0.39-0.41] | **R<sup>2</sup>:** 0.39 [0.35-0.42]<br>**MAE:** 0.43 [0.42-0.45] | **R<sup>2</sup>:** 0.46 [0.43-0.50]<br>**MAE:** 0.40 [0.38-0.41] | **R<sup>2</sup>:** 0.51 [0.47-0.54]<br>**MAE:** 0.37 [0.36-0.39] | **R<sup>2</sup>:** 0.43 [0.29-0.53]<br>**MAE:** 0.39 [0.36-0.43] | **R<sup>2</sup>:** 0.29 [0.01-0.47]<br>**MAE:** 0.43 [0.37-0.49] | **R<sup>2</sup>:** 0.57 [0.43-0.68]<br>**MAE:** 0.36 [0.32-0.41] | **R<sup>2</sup>:** 0.14 [-1.21-0.57]<br>**MAE:** 0.37 [0.31-0.44] | **R<sup>2</sup>:** 0.37 [0.29-0.44]<br>**MAE:** 0.35 [0.33-0.37] | **R<sup>2</sup>:** 0.16 [-0.74-0.38]<br>**MAE:** 0.43 [0.35-0.52] | **R<sup>2</sup>:** 0.36 [0.18-0.49]<br>**MAE:** 0.36 [0.33-0.39] | **R<sup>2</sup>:** 0.40 [0.29-0.48]<br>**MAE:** 0.34 [0.32-0.36] | — | — | — | — |

**Supplemental Figure 5.**
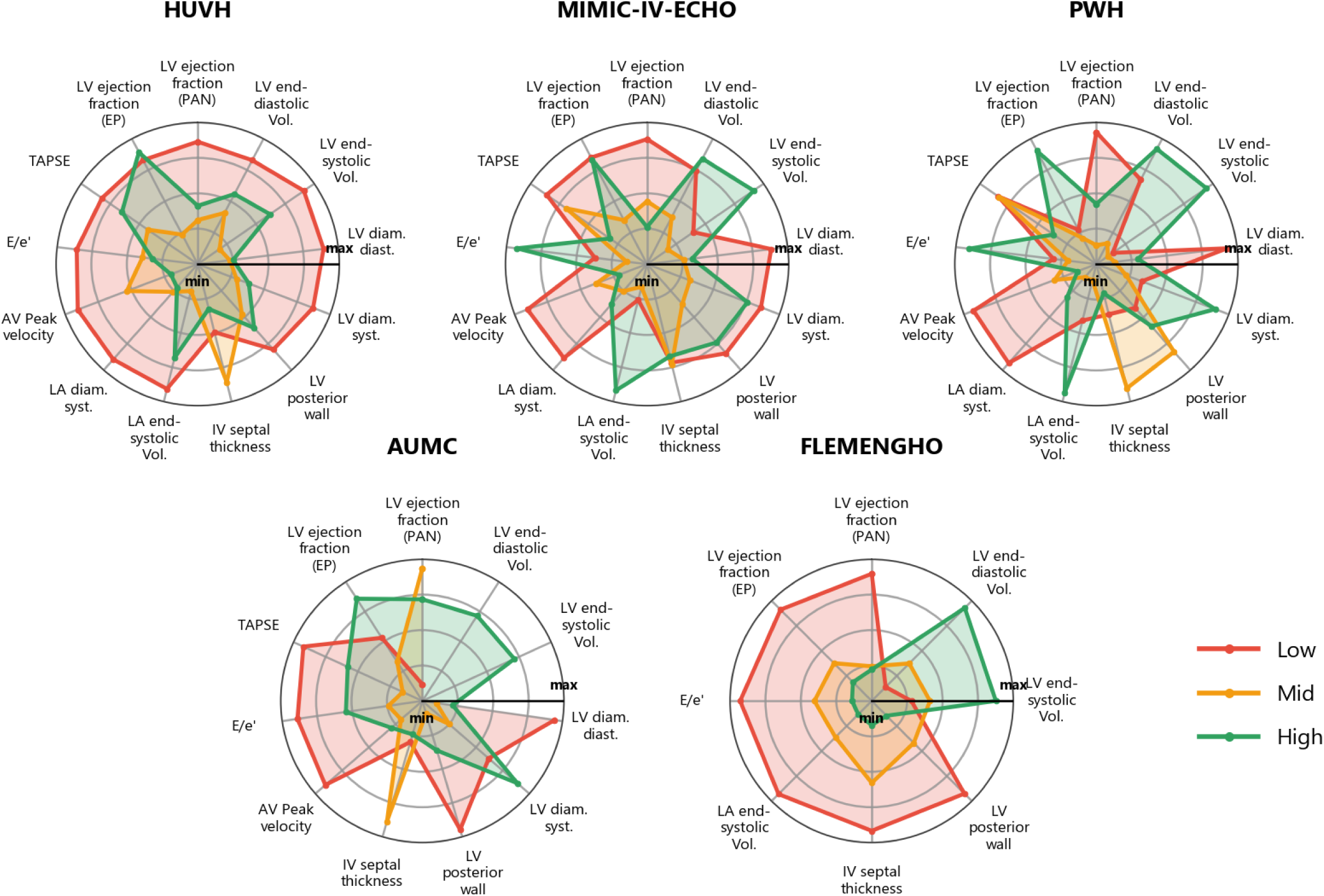
Measurements performance stratified by EchoQuality grade across cohorts, reported as mean absolute error (MAE). Values were normalized independently for each measurement within each cohort to enable cross-metric visual comparison.

**Supplemental Figure 6.**
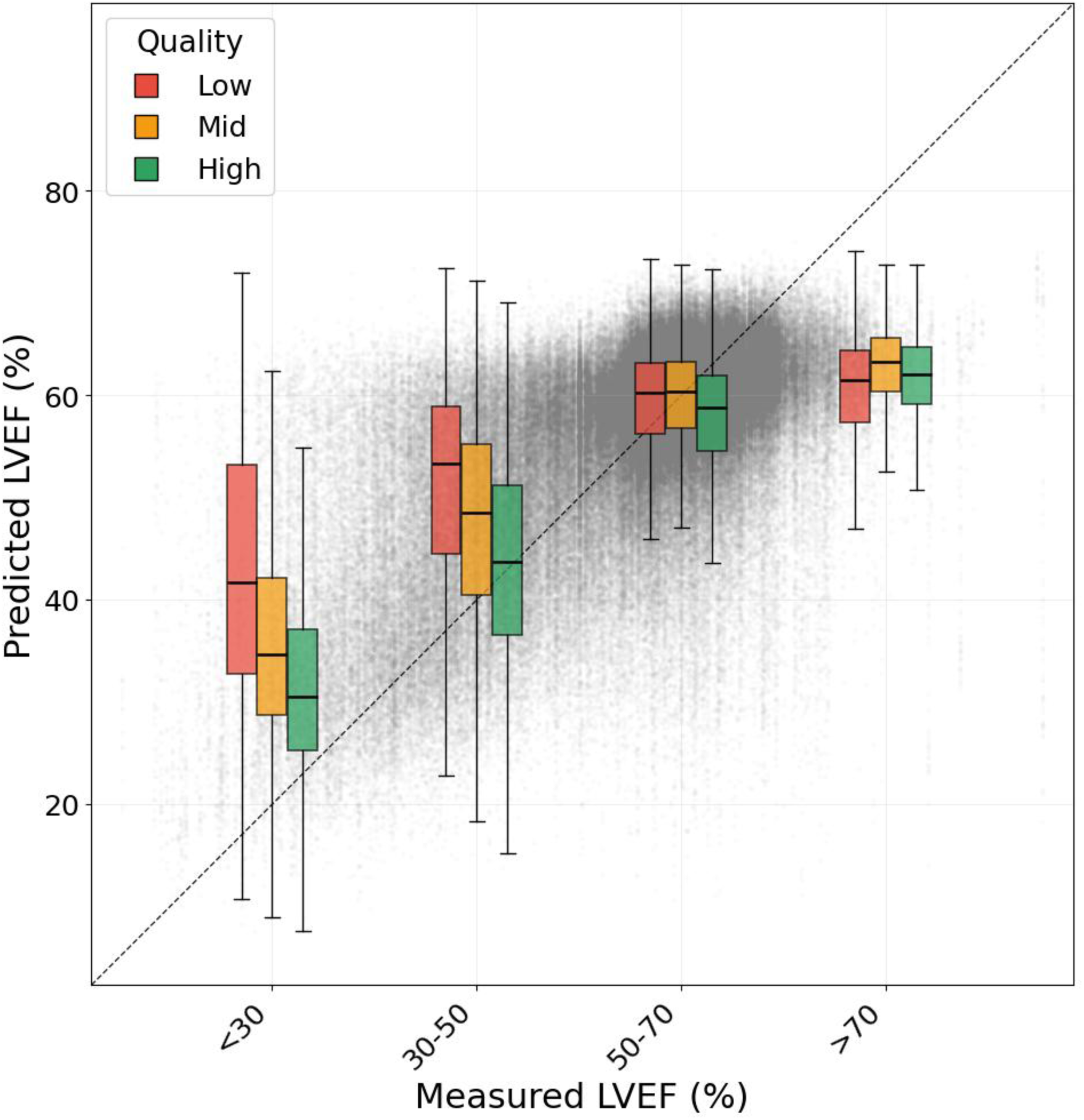
Distribution of video-level LVEF predictions from PanEcho in the HUVH cohort, stratified by EchoQuality grade.

**Supplemental Table 7.** Sensitivity and specificity of at least moderate dysfunction detections from echocardiographic measurements, stratified by EchoQuality grade across cohorts with 95% confidence intervals. Classifications were derived by applying published guideline thresholds to PanEcho and EchoPrime measurement outputs.

| Model | ALL |  |  |  | HUVH |  |  |  | MIMIC-IV-ECHO |  |  |  | AUMC |  |  |  | PWH |  |  |  | FLEMENGHO |  |  |  |
|---|---|---|---|---|---|---|---|---|---|---|---|---|---|---|---|---|---|---|---|---|---|---|---|---|
|  | All | Low | Mid | High | All | Low | Mid | High | All | Low | Mid | High | All | Low | Mid | High | All | Low | Mid | High | All | Low | Mid | High |
| ***Left ventricular dysfunction*** |  |  |  |  |  |  |  |  |  |  |  |  |  |  |  |  |  |  |  |  |  |  |  |  |
| *PanEcho* | **Sens:** 0.51 [0.49-0.53]<br>**Spec:** 0.99 [0.99-0.99] | **Sens:** 0.36 [0.32-0.42]<br>**Spec:** 1.00 [0.99-1.00] | **Sens:** 0.41 [0.38-0.45]<br>**Spec:** 0.99 [0.99-1.00] | **Sens:** 0.60 [0.57-0.62]<br>**Spec:** 0.98 [0.98-0.98] | **Sens:** 0.53 [0.50-0.55]<br>**Spec:** 0.99 [0.99-0.99] | **Sens:** 0.41 [0.35-0.47]<br>**Spec:** 0.99 [0.99-1.00] | **Sens:** 0.44 [0.40-0.48]<br>**Spec:** 0.99 [0.99-0.99] | **Sens:** 0.62 [0.58-0.65]<br>**Spec:** 0.98 [0.98-0.98] | **Sens:** 0.42 [0.37-0.47]<br>**Spec:** 0.99 [0.99-1.00] | **Sens:** 0.19 [0.11-0.28]<br>**Spec:** 1.00 [1.00-1.00] | **Sens:** 0.23 [0.14-0.32]<br>**Spec:** 1.00 [0.99-1.00] | **Sens:** 0.58 [0.52-0.64]<br>**Spec:** 0.98 [0.97-0.99] | **Sens:** 0.35 [0.23-0.50]<br>**Spec:** 0.99 [0.98-1.00] | — | — | **Sens:** 0.41 [0.23-0.59]<br>**Spec:** 0.98 [0.95-1.00] | **Sens:** 0.56 [0.49-0.63]<br>**Spec:** 0.99 [0.98-1.00] | — | **Sens:** 0.50 [0.30-0.71]<br>**Spec:** 1.00 [0.99-1.00] | **Sens:** 0.58 [0.50-0.65]<br>**Spec:** 0.98 [0.97-0.99] | — | — | — | — |
| *EchoPrime* | **Sens:** 0.05 [0.04-0.06]<br>**Spec:** 1.00 [1.00-1.00] | **Sens:** 0.02 [0.01-0.05]<br>**Spec:** 1.00 [1.00-1.00] | **Sens:** 0.03 [0.02-0.04]<br>**Spec:** 1.00 [1.00-1.00] | **Sens:** 0.06 [0.05-0.08]<br>**Spec:** 1.00 [1.00-1.00] | **Sens:** 0.04 [0.03-0.05]<br>**Spec:** 1.00 [1.00-1.00] | **Sens:** 0.03 [0.01-0.05]<br>**Spec:** 1.00 [1.00-1.00] | **Sens:** 0.02 [0.01-0.04]<br>**Spec:** 1.00 [1.00-1.00] | **Sens:** 0.06 [0.04-0.07]<br>**Spec:** 1.00 [1.00-1.00] | **Sens:** 0.09 [0.06-0.12]<br>**Spec:** 1.00 [1.00-1.00] | **Sens:** 0.02 [0.00-0.06]<br>**Spec:** 1.00 [1.00-1.00] | **Sens:** 0.06 [0.01-0.11]<br>**Spec:** 1.00 [1.00-1.00] | **Sens:** 0.12 [0.08-0.17]<br>**Spec:** 0.99 [0.99-1.00] | **Sens:** 0.04 [0.00-0.11]<br>**Spec:** 1.00 [0.99-1.00] | — | — | **Sens:** 0.04 [0.00-0.13]<br>**Spec:** 0.99 [0.97-1.00] | **Sens:** 0.02 [0.00-0.05]<br>**Spec:** 1.00 [1.00-1.00] | — | **Sens:** 0.00 [0.00-0.00]<br>**Spec:** 1.00 [1.00-1.00] | **Sens:** 0.02 [0.01-0.05]<br>**Spec:** 1.00 [1.00-1.00] | — | — | — | — |
| ***Aortic stenosis*** |  |  |  |  |  |  |  |  |  |  |  |  |  |  |  |  |  |  |  |  |  |  |  |  |
| *PanEcho* | **Sens:** 0.26 [0.24-0.28]<br>**Spec:** 0.99 [0.99-0.99] | **Sens:** 0.23 [0.20-0.27]<br>**Spec:** 0.99 [0.99-0.99] | **Sens:** 0.27 [0.25-0.29]<br>**Spec:** 0.99 [0.99-0.99] | **Sens:** 0.26 [0.24-0.29]<br>**Spec:** 1.00 [0.99-1.00] | **Sens:** 0.28 [0.26-0.30]<br>**Spec:** 0.99 [0.99-0.99] | **Sens:** 0.28 [0.24-0.32]<br>**Spec:** 0.99 [0.98-0.99] | **Sens:** 0.28 [0.25-0.31]<br>**Spec:** 0.99 [0.99-0.99] | **Sens:** 0.28 [0.25-0.31]<br>**Spec:** 0.99 [0.99-1.00] | **Sens:** 0.14 [0.11-0.17]<br>**Spec:** 1.00 [1.00-1.00] | **Sens:** 0.10 [0.05-0.16]<br>**Spec:** 1.00 [0.99-1.00] | **Sens:** 0.14 [0.09-0.20]<br>**Spec:** 1.00 [0.99-1.00] | **Sens:** 0.19 [0.12-0.26]<br>**Spec:** 1.00 [1.00-1.00] | **Sens:** 0.38 [0.16-0.59]<br>**Spec:** 0.98 [0.96-0.99] | — | — | — | **Sens:** 0.18 [0.06-0.31]<br>**Spec:** 1.00 [1.00-1.00] | — | — | **Sens:** 0.18 [0.03-0.37]<br>**Spec:** 1.00 [1.00-1.00] | — | — | — | — |
| ***Left ventricular dilation*** |  |  |  |  |  |  |  |  |  |  |  |  |  |  |  |  |  |  |  |  |  |  |  |  |
| *PanEcho* | **Sens:** 0.45 [0.43-0.47]<br>**Spec:** 0.95 [0.95-0.95] | **Sens:** 0.31 [0.26-0.38]<br>**Spec:** 0.98 [0.97-0.98] | **Sens:** 0.35 [0.30-0.39]<br>**Spec:** 0.97 [0.96-0.97] | **Sens:** 0.52 [0.49-0.55]<br>**Spec:** 0.92 [0.91-0.92] | **Sens:** 0.50 [0.47-0.53]<br>**Spec:** 0.93 [0.93-0.94] | **Sens:** 0.38 [0.31-0.45]<br>**Spec:** 0.96 [0.95-0.97] | **Sens:** 0.41 [0.36-0.46]<br>**Spec:** 0.95 [0.95-0.96] | **Sens:** 0.57 [0.53-0.60]<br>**Spec:** 0.89 [0.89-0.90] | **Sens:** 0.19 [0.14-0.23]<br>**Spec:** 1.00 [0.99-1.00] | **Sens:** 0.13 [0.04-0.24]<br>**Spec:** 1.00 [0.99-1.00] | **Sens:** 0.03 [0.00-0.08]<br>**Spec:** 1.00 [0.99-1.00] | **Sens:** 0.26 [0.19-0.32]<br>**Spec:** 0.99 [0.98-1.00] | **Sens:** 0.44 [0.26-0.64]<br>**Spec:** 0.95 [0.90-0.99] | — | — | — | **Sens:** 0.53 [0.47-0.59]<br>**Spec:** 0.98 [0.97-0.99] | — | **Sens:** 0.32 [0.17-0.48]<br>**Spec:** 0.99 [0.97-1.00] | **Sens:** 0.58 [0.51-0.65]<br>**Spec:** 0.97 [0.96-0.98] | — | — | — | — |
| ***Abnormal left ventricular thickness*** |  |  |  |  |  |  |  |  |  |  |  |  |  |  |  |  |  |  |  |  |  |  |  |  |
| *PanEcho* | **Sens:** 0.11 [0.10-0.12]<br>**Spec:** 1.00 [1.00-1.00] | **Sens:** 0.05 [0.04-0.06]<br>**Spec:** 1.00 [1.00-1.00] | **Sens:** 0.10 [0.09-0.11]<br>**Spec:** 1.00 [1.00-1.00] | **Sens:** 0.16 [0.15-0.18]<br>**Spec:** 1.00 [1.00-1.00] | **Sens:** 0.12 [0.12-0.13]<br>**Spec:** 1.00 [1.00-1.00] | **Sens:** 0.05 [0.04-0.07]<br>**Spec:** 1.00 [0.99-1.00] | **Sens:** 0.11 [0.10-0.12]<br>**Spec:** 1.00 [1.00-1.00] | **Sens:** 0.18 [0.16-0.20]<br>**Spec:** 1.00 [1.00-1.00] | **Sens:** 0.05 [0.04-0.06]<br>**Spec:** 1.00 [1.00-1.00] | **Sens:** 0.02 [0.01-0.03]<br>**Spec:** 1.00 [1.00-1.00] | **Sens:** 0.04 [0.03-0.06]<br>**Spec:** 1.00 [1.00-1.00] | **Sens:** 0.09 [0.06-0.12]<br>**Spec:** 1.00 [1.00-1.00] | **Sens:** 0.20 [0.07-0.35]<br>**Spec:** 1.00 [0.99-1.00] | — | — | — | **Sens:** 0.13 [0.09-0.18]<br>**Spec:** 1.00 [1.00-1.00] | — | **Sens:** 0.15 [0.07-0.23]<br>**Spec:** 1.00 [0.99-1.00] | **Sens:** 0.13 [0.07-0.19]<br>**Spec:** 1.00 [1.00-1.00] | — | — | — | — |
| ***Left atrium dilation*** |  |  |  |  |  |  |  |  |  |  |  |  |  |  |  |  |  |  |  |  |  |  |  |  |
| *PanEcho* | **Sens:** 0.42 [0.41-0.44]<br>**Spec:** 0.96 [0.96-0.97] | **Sens:** 0.29 [0.26-0.32]<br>**Spec:** 0.98 [0.98-0.99] | **Sens:** 0.36 [0.34-0.38]<br>**Spec:** 0.97 [0.97-0.98] | **Sens:** 0.52 [0.50-0.54]<br>**Spec:** 0.94 [0.93-0.94] | **Sens:** 0.47 [0.45-0.48]<br>**Spec:** 0.95 [0.94-0.96] | **Sens:** 0.35 [0.32-0.39]<br>**Spec:** 0.96 [0.95-0.97] | **Sens:** 0.39 [0.37-0.41]<br>**Spec:** 0.96 [0.95-0.97] | **Sens:** 0.57 [0.55-0.59]<br>**Spec:** 0.93 [0.92-0.94] | **Sens:** 0.25 [0.22-0.27]<br>**Spec:** 0.97 [0.97-0.98] | **Sens:** 0.12 [0.08-0.16]<br>**Spec:** 0.99 [0.98-1.00] | **Sens:** 0.16 [0.12-0.20]<br>**Spec:** 0.99 [0.98-0.99] | **Sens:** 0.35 [0.31-0.38]<br>**Spec:** 0.93 [0.91-0.94] | — | — | — | — | **Sens:** 0.63 [0.58-0.68]<br>**Spec:** 0.92 [0.90-0.94] | — | **Sens:** 0.55 [0.44-0.66]<br>**Spec:** 0.96 [0.93-0.98] | **Sens:** 0.63 [0.57-0.69]<br>**Spec:** 0.89 [0.87-0.92] | — | — | — | — |

**Supplemental Table 8.**
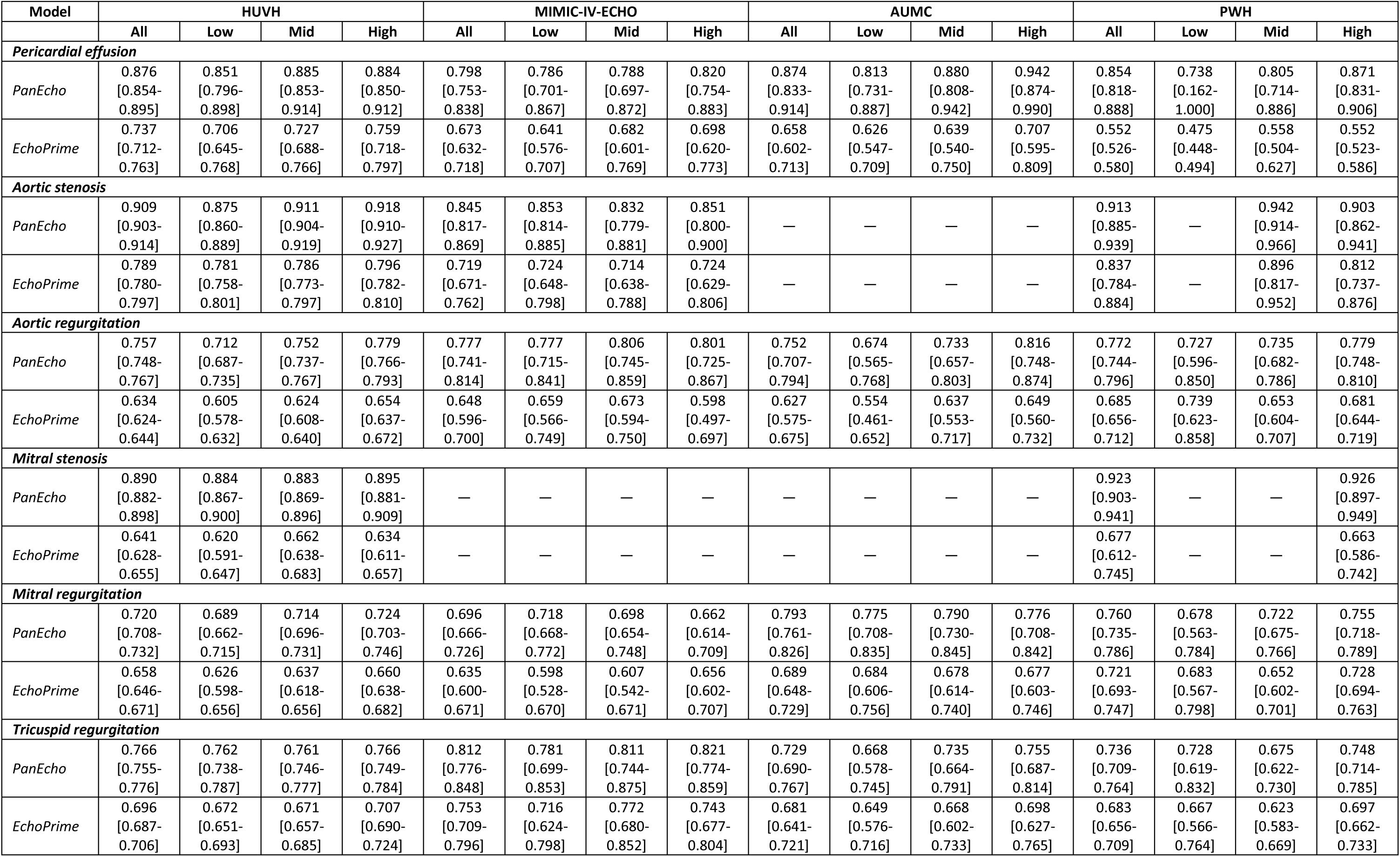
Disease classification performance stratified by EchoQuality grade across cohorts, reported as area under the receiver operating characteristic curve (AUROC).

| Model | HUVH |  |  |  | MIMIC-IV-ECHO |  |  |  | AUMC |  |  |  | PWH |  |  |  |
|---|---|---|---|---|---|---|---|---|---|---|---|---|---|---|---|---|
|  | All | Low | Mid | High | All | Low | Mid | High | All | Low | Mid | High | All | Low | Mid | High |
| ***Pericardial effusion*** |  |  |  |  |  |  |  |  |  |  |  |  |  |  |  |  |
| *PanEcho* | 0.876 [0.854-0.895] | 0.851 [0.796-0.898] | 0.885 [0.853-0.914] | 0.884 [0.850-0.912] | 0.798 [0.753-0.838] | 0.786 [0.701-0.867] | 0.788 [0.697-0.872] | 0.820 [0.754-0.883] | 0.874 [0.833-0.914] | 0.813 [0.731-0.887] | 0.880 [0.808-0.942] | 0.942 [0.874-0.990] | 0.854 [0.818-0.888] | 0.738 [0.162-1.000] | 0.805 [0.714-0.886] | 0.871 [0.831-0.906] |
| *EchoPrime* | 0.737 [0.712-0.763] | 0.706 [0.645-0.768] | 0.727 [0.688-0.766] | 0.759 [0.718-0.797] | 0.673 [0.632-0.718] | 0.641 [0.576-0.707] | 0.682 [0.601-0.769] | 0.698 [0.620-0.773] | 0.658 [0.602-0.713] | 0.626 [0.547-0.709] | 0.639 [0.540-0.750] | 0.707 [0.595-0.809] | 0.552 [0.526-0.580] | 0.475 [0.448-0.494] | 0.558 [0.504-0.627] | 0.552 [0.523-0.586] |
| ***Aortic stenosis*** |  |  |  |  |  |  |  |  |  |  |  |  |  |  |  |  |
| *PanEcho* | 0.909 [0.903-0.914] | 0.875 [0.860-0.889] | 0.911 [0.904-0.919] | 0.918 [0.910-0.927] | 0.845 [0.817-0.869] | 0.853 [0.814-0.885] | 0.832 [0.779-0.881] | 0.851 [0.800-0.900] | — | — | — | — | 0.913 [0.885-0.939] | — | 0.942 [0.914-0.966] | 0.903 [0.862-0.941] |
| *EchoPrime* | 0.789 [0.780-0.797] | 0.781 [0.758-0.801] | 0.786 [0.773-0.797] | 0.796 [0.782-0.810] | 0.719 [0.671-0.762] | 0.724 [0.648-0.798] | 0.714 [0.638-0.788] | 0.724 [0.629-0.806] | — | — | — | — | 0.837 [0.784-0.884] | — | 0.896 [0.817-0.952] | 0.812 [0.737-0.876] |
| ***Aortic regurgitation*** |  |  |  |  |  |  |  |  |  |  |  |  |  |  |  |  |
| *PanEcho* | 0.757 [0.748-0.767] | 0.712 [0.687-0.735] | 0.752 [0.737-0.767] | 0.779 [0.766-0.793] | 0.777 [0.741-0.814] | 0.777 [0.715-0.841] | 0.806 [0.745-0.859] | 0.801 [0.725-0.867] | 0.752 [0.707-0.794] | 0.674 [0.565-0.768] | 0.733 [0.657-0.803] | 0.816 [0.748-0.874] | 0.772 [0.744-0.796] | 0.727 [0.596-0.850] | 0.735 [0.682-0.786] | 0.779 [0.748-0.810] |
| *EchoPrime* | 0.634 [0.624-0.644] | 0.605 [0.578-0.632] | 0.624 [0.608-0.640] | 0.654 [0.637-0.672] | 0.648 [0.596-0.700] | 0.659 [0.566-0.749] | 0.673 [0.594-0.750] | 0.598 [0.497-0.697] | 0.627 [0.575-0.675] | 0.554 [0.461-0.652] | 0.637 [0.553-0.717] | 0.649 [0.560-0.732] | 0.685 [0.656-0.712] | 0.739 [0.623-0.858] | 0.653 [0.604-0.707] | 0.681 [0.644-0.719] |
| ***Mitral stenosis*** |  |  |  |  |  |  |  |  |  |  |  |  |  |  |  |  |
| *PanEcho* | 0.890 [0.882-0.898] | 0.884 [0.867-0.900] | 0.883 [0.869-0.896] | 0.895 [0.881-0.909] | — | — | — | — | — | — | — | — | 0.923 [0.903-0.941] | — | — | 0.926 [0.897-0.949] |
| *EchoPrime* | 0.641 [0.628-0.655] | 0.620 [0.591-0.647] | 0.662 [0.638-0.683] | 0.634 [0.611-0.657] | — | — | — | — | — | — | — | — | 0.677 [0.612-0.745] | — | — | 0.663 [0.586-0.742] |
| ***Mitral regurgitation*** |  |  |  |  |  |  |  |  |  |  |  |  |  |  |  |  |
| *PanEcho* | 0.720 [0.708-0.732] | 0.689 [0.662-0.715] | 0.714 [0.696-0.731] | 0.724 [0.703-0.746] | 0.696 [0.666-0.726] | 0.718 [0.668-0.772] | 0.698 [0.654-0.748] | 0.662 [0.614-0.709] | 0.793 [0.761-0.826] | 0.775 [0.708-0.835] | 0.790 [0.730-0.845] | 0.776 [0.708-0.842] | 0.760 [0.735-0.786] | 0.678 [0.563-0.784] | 0.722 [0.675-0.766] | 0.755 [0.718-0.789] |
| *EchoPrime* | 0.658 [0.646-0.671] | 0.626 [0.598-0.656] | 0.637 [0.618-0.656] | 0.660 [0.638-0.682] | 0.635 [0.600-0.671] | 0.598 [0.528-0.670] | 0.607 [0.542-0.671] | 0.656 [0.602-0.707] | 0.689 [0.648-0.729] | 0.684 [0.606-0.756] | 0.678 [0.614-0.740] | 0.677 [0.603-0.746] | 0.721 [0.693-0.747] | 0.683 [0.567-0.798] | 0.652 [0.602-0.701] | 0.728 [0.694-0.763] |
| ***Tricuspid regurgitation*** |  |  |  |  |  |  |  |  |  |  |  |  |  |  |  |  |
| *PanEcho* | 0.766 [0.755-0.776] | 0.762 [0.738-0.787] | 0.761 [0.746-0.777] | 0.766 [0.749-0.784] | 0.812 [0.776-0.848] | 0.781 [0.699-0.853] | 0.811 [0.744-0.875] | 0.821 [0.774-0.859] | 0.729 [0.690-0.767] | 0.668 [0.578-0.745] | 0.735 [0.664-0.791] | 0.755 [0.687-0.814] | 0.736 [0.709-0.764] | 0.728 [0.619-0.832] | 0.675 [0.622-0.730] | 0.748 [0.714-0.785] |
| *EchoPrime* | 0.696 [0.687-0.706] | 0.672 [0.651-0.693] | 0.671 [0.657-0.685] | 0.707 [0.690-0.724] | 0.753 [0.709-0.796] | 0.716 [0.624-0.798] | 0.772 [0.680-0.852] | 0.743 [0.677-0.804] | 0.681 [0.641-0.721] | 0.649 [0.576-0.716] | 0.668 [0.602-0.733] | 0.698 [0.627-0.765] | 0.683 [0.656-0.709] | 0.667 [0.566-0.764] | 0.623 [0.583-0.669] | 0.697 [0.662-0.733] |

**Supplemental Figure 7.**
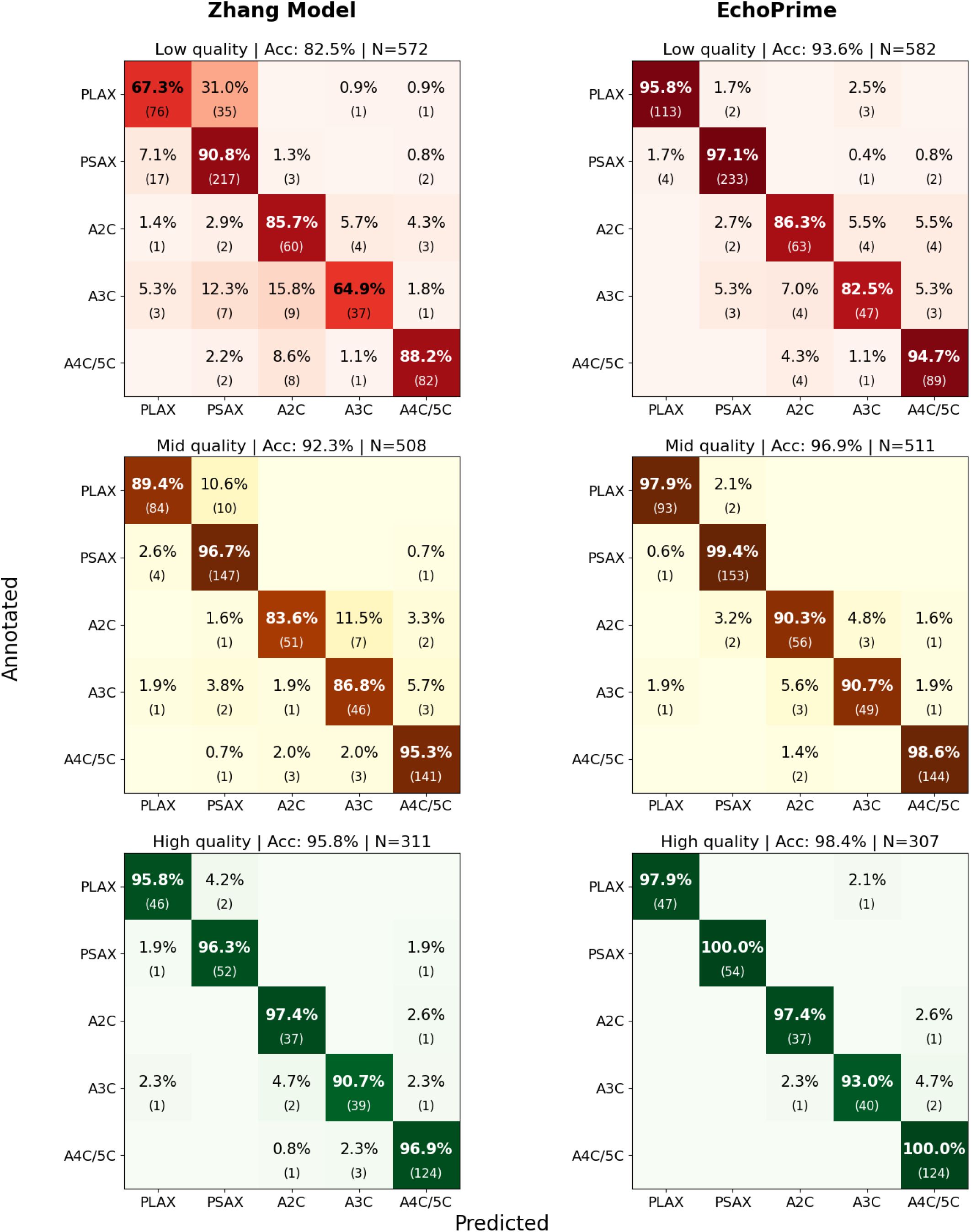
Confusion matrices for view identification performance of the Zhang 17 and EchoPrime 18 models stratified by EchoQuality grade (low, mid, high) in the HUVH-view

**Supplemental Table 9.** Comparison of baseline and quality-aware aggregation performance across measurements and detection of moderate dysfunctions, stratified by cohort. Baseline performance reflects uniform averaging of video-level predictions to study level. Quality-aware performance reflects weighted aggregation and filtering using the linear EchoQuality score.

|  | ALL |  | HUVH |  | MIMIC-IV-ECHO |  | AUMC |  | PWH |  | FLEMENGHO |  |
|---|---|---|---|---|---|---|---|---|---|---|---|---|
|  | Baseline | Quality-aware | Baseline | Quality-aware | Baseline | Quality-aware | Baseline | Quality-aware | Baseline | Quality-aware | Baseline | Quality-aware |
| ***Measurements (R<sup>2</sup> & MAE)*** |  |  |  |  |  |  |  |  |  |  |  |  |
| *Left ventricle ejection fraction* | — |  | R²: 0.63 [0.62-0.64] MAE: 5.64 [5.56-5.72] | R²: 0.65 [0.64-0.66] MAE: 5.52 [5.45-5.60] | R²: 0.48 [0.46-0.51] MAE: 7.63 [7.44-7.82] | R²: 0.49 [0.47-0.52] MAE: 7.57 [7.36-7.77] | R²: 0.45 [0.35-0.54] MAE: 6.15 [5.65-6.62] | R²: 0.46 [0.34-0.55] MAE: 6.14 [5.65-6.64] | R²: 0.59 [0.54-0.63] MAE: 5.60 [5.33-5.86] | R²: 0.61 [0.57-0.66] MAE: 5.42 [5.18-5.68] | R²: 0.15 [0.10-0.20] MAE: 4.36 [4.18-4.56] | R²: 0.14 [0.08-0.20] MAE: 4.33 [4.14-4.52] |
| *Left ventricle end-diastolic volume* | — |  | R²: 0.41 [0.39-0.42] MAE: 26.45 [26.12-26.79] | R²: 0.42 [0.41-0.44] MAE: 25.89 [25.56-26.23] | R²: 0.39 [0.36-0.41] MAE: 24.90 [24.08-25.67] | R²: 0.41 [0.38-0.44] MAE: 24.31 [23.52-25.15] | R²: 0.61 [0.54-0.69] MAE: 23.97 [19.10-30.79] | R²: 0.60 [0.50-0.72] MAE: 23.08 [17.77-29.80] | R²: 0.63 [0.61-0.66] MAE: 20.76 [19.71-21.83] | R²: 0.66 [0.63-0.69] MAE: 20.24 [19.26-21.34] | R²: 0.12 [0.05-0.18] MAE: 18.85 [17.99-19.73] | R²: 0.04 [-0.04-0.11] MAE: 19.56 [18.67-20.42] |
| *Left ventricle end-systolic volume* | — |  | R²: 0.65 [0.63-0.67] MAE: 16.64 [16.15-17.18] | R²: 0.67 [0.66-0.69] MAE: 16.28 [15.73-16.80] | R²: 0.58 [0.56-0.61] MAE: 13.38 [12.87-13.91] | R²: 0.62 [0.59-0.64] MAE: 12.91 [12.42-13.38] | R²: 0.56 [0.49-0.70] MAE: 17.34 [12.66-22.87] | R²: 0.57 [0.47-0.72] MAE: 16.92 [12.32-23.31] | R²: 0.70 [0.67-0.73] MAE: 13.50 [12.65-14.38] | R²: 0.74 [0.71-0.76] MAE: 12.95 [12.18-13.74] | R²: 0.22 [0.14-0.30] MAE: 8.28 [7.86-8.70] | R²: 0.19 [0.11-0.26] MAE: 8.46 [8.03-8.87] |
| *Interventricular septal thickness* | — |  | R²: 0.19 [0.17-0.20] MAE: 0.21 [0.21-0.21] | R²: 0.20 [0.18-0.22] MAE: 0.21 [0.21-0.22] | R²: 0.28 [0.26-0.30] MAE: 0.16 [0.15-0.16] | R²: 0.29 [0.27-0.32] MAE: 0.16 [0.15-0.16] | R²: 0.22 [0.12-0.43] MAE: 0.17 [0.15-0.19] | R²: 0.21 [0.09-0.45] MAE: 0.16 [0.15-0.19] | R²: 0.40 [0.36-0.45] MAE: 0.15 [0.15-0.16] | R²: 0.41 [0.36-0.46] MAE: 0.15 [0.14-0.16] | R²: 0.29 [0.20-0.38] MAE: 0.10 [0.10-0.11] | R²: 0.40 [0.30-0.48] MAE: 0.09 [0.08-0.10] |
| *Left ventricle posterior wall thickness* | — |  | R²: 0.18 [0.13-0.25] MAE: 0.15 [0.15-0.15] | R²: 0.18 [0.13-0.25] MAE: 0.15 [0.15-0.15] | R²: 0.15 [0.13-0.18] MAE: 0.16 [0.16-0.16] | R²: 0.17 [0.15-0.20] MAE: 0.16 [0.15-0.16] | R²: 0.08 [0.04-0.23] MAE: 0.20 [0.16-0.26] | R²: 0.10 [0.06-0.29] MAE: 0.20 [0.15-0.26] | R²: 0.30 [0.26-0.35] MAE: 0.14 [0.14-0.15] | R²: 0.31 [0.26-0.35] MAE: 0.14 [0.14-0.15] | R²: 0.15 [0.03-0.24] MAE: 0.09 [0.09-0.10] | R²: 0.32 [0.23-0.41] MAE: 0.08 [0.08-0.09] |
| *Left ventricle interior diameter systole* | — |  | R²: 0.56 [0.55-0.57] MAE: 0.48 [0.47-0.49] | R²: 0.58 [0.57-0.60] MAE: 0.47 [0.46-0.48] | R²: 0.56 [0.54-0.57] MAE: 0.43 [0.42-0.44] | R²: 0.61 [0.60-0.63] MAE: 0.41 [0.40-0.42] | R²: 0.50 [0.39-0.58] MAE: 0.43 [0.39-0.47] | R²: 0.58 [0.48-0.67] MAE: 0.39 [0.35-0.44] | R²: 0.63 [0.61-0.65] MAE: 0.44 [0.41-0.46] | R²: 0.67 [0.64-0.70] MAE: 0.41 [0.40-0.43] | — | — |
| *Left ventricle interior diameter diastole* | — |  | R²: 0.50 [0.49-0.51] MAE: 0.45 [0.45-0.46] | R²: 0.53 [0.52-0.54] MAE: 0.44 [0.43-0.45] | R²: 0.45 [0.43-0.46] MAE: 0.46 [0.45-0.47] | R²: 0.49 [0.48-0.51] MAE: 0.44 [0.43-0.45] | R²: 0.26 [0.08-0.56] MAE: 0.46 [0.39-0.55] | R²: 0.26 [0.05-0.61] MAE: 0.44 [0.37-0.55] | R²: 0.58 [0.55-0.60] MAE: 0.40 [0.38-0.42] | R²: 0.61 [0.58-0.64] MAE: 0.38 [0.37-0.40] | — | — |
| *E/E' ratio* | — |  | R²: 0.52 [0.50-0.53] MAE: 3.02 [2.94-3.09] | R²: 0.53 [0.51-0.54] MAE: 2.96 [2.88-3.03] | R²: 0.50 [0.48-0.51] MAE: 3.02 [2.92-3.12] | R²: 0.51 [0.49-0.53] MAE: 3.01 [2.89-3.11] | R²: 0.60 [0.53-0.65] MAE: 1.88 [1.67-2.11] | R²: 0.62 [0.55-0.68] MAE: 1.81 [1.60-2.03] | R²: 0.42 [0.35-0.48] MAE: 3.06 [2.88-3.24] | R²: 0.42 [0.35-0.48] MAE: 3.06 [2.88-3.25] | R²: -0.42 [-0.77--0.13] MAE: 2.53 [2.47-2.61] | R²: -0.20 [-0.53-0.02] MAE: 2.26 [2.19-2.33] |
| *TAPSE* | — |  | R²: 0.32 [0.30-0.33] MAE: 0.29 [0.29-0.29] | R²: 0.32 [0.30-0.33] MAE: 0.29 [0.29-0.29] | R²: 0.14 [0.10-0.18] MAE: 0.40 [0.38-0.41] | R²: 0.14 [0.09-0.19] MAE: 0.40 [0.38-0.41] | R²: 0.29 [0.22-0.36] MAE: 0.36 [0.33-0.38] | R²: 0.31 [0.22-0.38] MAE: 0.35 [0.33-0.37] | R²: 0.08 [0.02-0.14] MAE: 0.31 [0.30-0.33] | R²: 0.08 [0.01-0.14] MAE: 0.32 [0.30-0.33] | — | — |
| *Left atrium internal diameter systole* | — |  | R²: 0.49 [0.48-0.51] | R²: 0.50 [0.48-0.52] | R²: 0.45 [0.44-0.47] | R²: 0.48 [0.46-0.50] | — | — | R²: 0.48 [0.43-0.52] | R²: 0.48 [0.43-0.53] | — | — |

**Supplemental Figure 8.**
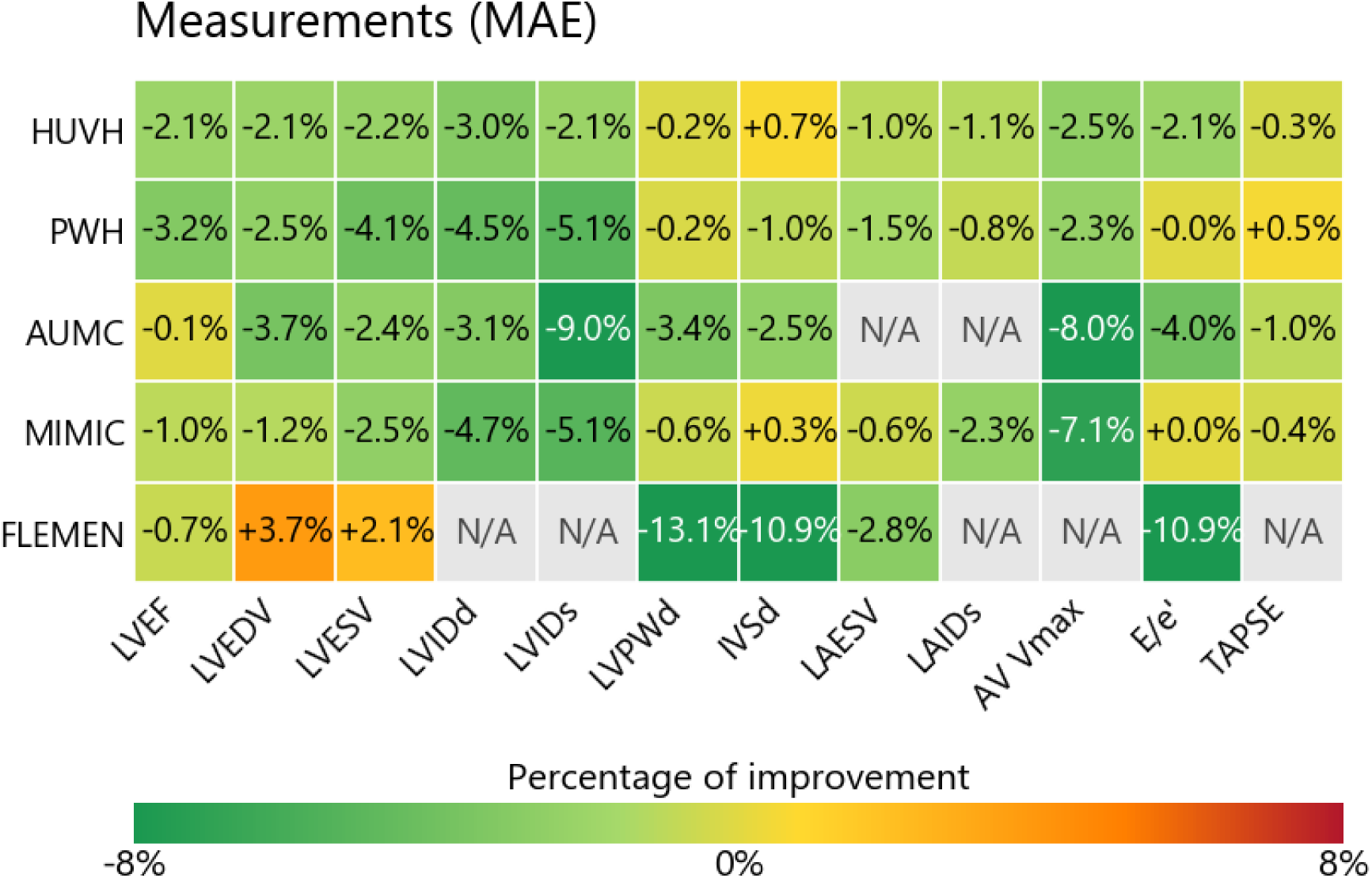
Impact of quality-aware aggregation on echocardiographic measurement, quantified as the change in mean absolute error (MAE) relative to baseline uniform aggregation. N/A refers to tests whose number of observations was not large enough to compute robust statistics or whose labels were not available.

**Supplemental Figure 9.**
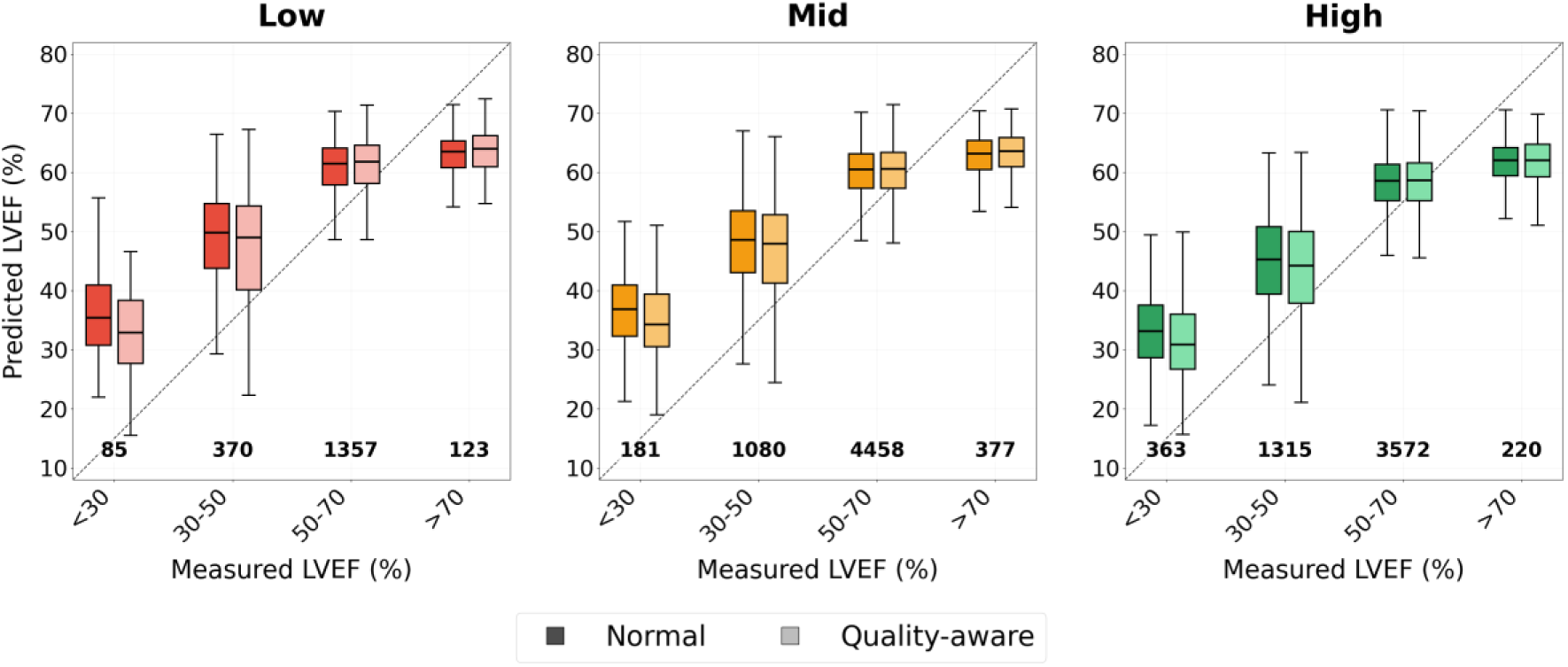
Comparison of quality-aware aggregation of LVEF against uniform averaging as the study-level aggregation baseline.

